# Metformin on the Presence of COVID-19 Symptoms Over 6 Months: The ACTIV-6 Randomized Clinical Trial

**DOI:** 10.1101/2025.08.08.25333305

**Authors:** Carolyn T. Bramante, Thomas G. Stewart, David R. Boulware, Matthew W. McCarthy, Yue Gao, Russell L. Rothman, Ahmad Mourad, Florence Thicklin, Jonathan B. Cohen, Idania T. Garcia del Sol, Nirav S. Shah, Manisha Mehta, Orlando Quintero Cardona, Jake Scott, Adit A. Ginde, Mario Castro, Dushyantha Jayaweera, Mark Sulkowski, Nina Gentile, Kathleen McTigue, G. Michael Felker, Sean Collins, Sarah E. Dunsmore, Stacey J. Adam, Christopher J. Lindsell, Adrian F. Hernandez, Susanna Naggie, the Accelerating COVID-19 Therapeutic Interventions and Vaccines (ACTIV)-6 Study Group and Investigators

**Affiliations:** Department of Medicine, University of Minnesota Medical School, Minneapolis, MN; School of Data Science, University of Virginia, Charlottesville, VA; Weill Cornell Medicine, New York, NY; Vanderbilt University Medical Center, Nashville, TN; Department of Medicine, Duke University School of Medicine, Durham, NC; Duke Clinical Research Institute, Duke University School of Medicine, Durham, NC; Stakeholder Advisory Committee, Pittsburgh, PA; Jadestone Clinical Research, LLC, Silver Spring, MD; L&A Morales Healthcare, Inc., Miami, FL; Endeavor Health, Evanston, IL; GFC of Southeastern Michigan, PC, Detroit, MI; Stanford University School of Medicine, Department of Medicine, Division of Infectious Diseases, Stanford, CA; Department of Emergency Medicine, University of Colorado School of Medicine, Aurora, CO; Division of Pulmonary, Critical Care and Sleep Medicine, University of Kansas Medical Center, Kansas City, KS; Department of Medicine, Miller School of Medicine, University of Miami, Miami, FL; Division of Infectious Diseases, Johns Hopkins University, Baltimore, MD; Department of Emergency Medicine, Lewis Katz School of Medicine at Temple University, Philadelphia, PA; Department of Medicine, University of Pittsburgh Medical Center, Pittsburgh, PA; Veterans Affairs Tennessee Valley Healthcare System, Geriatric Research, Education and Clinical Center (GRECC), Nashville, TN; National Center for Advancing Translational Sciences, Bethesda, MD; Foundation for the National Institutes of Health, Bethesda, MD

## Abstract

**Background:** The effect of metformin on preventing long-term COVID-19 symptoms among low-risk adults has not been studied. The objective of this study was to Assess metformin compared with placebo during acute SARS-CoV-2 infection on the presence of COVID-19 symptoms 180 days later.

**Methods:** The ACTIV-6 platform evaluated repurposed medications for mild to moderate COVID-19. Between September 19, 2023 and May 1, 2024, 2983 outpatient adults ≥30 years with confirmed SARS-CoV-2 infection and ≥2 COVID-19 symptoms for ≤7 days were included from 90 sites. Participants were randomized to metformin (titrated to 1500 mg daily) or placebo for 14 days. Post-acute sequelae of SARS-CoV-2 or death (PASCD) was ascertained by asking whether participants had symptoms they attributed to COVID-19 on day 180. Secondary outcomes included clinician diagnosis of long COVID. For the primary outcome, the single-sided threshold for efficacy was 0.975.

**Results:** Among 2983 participants, the median age was 47 years (interquartile range [IQR] 38–57); 63% were female; 47% Hispanic/Latino; 83% reported ≥1 prior COVID-19 infections or SARS-CoV-2 vaccines. There were no deaths. Overall, 96 (3.2%) reported COVID-19 symptoms on day 90, 101 (3.4%) on day 120, and 79 (2.6%) on day 180. The covariate-adjusted risk of PASCD on day 180 was lower in the metformin group (−0.008; 95% credible interval [CrI] −0.022 to 0.006; posterior probability of efficacy [PPE] 0.83), compared with the placebo group with an adjusted risk ratio of 0.79 (95% CrI 0.474 to 1.230). The risk of clinician diagnosis of long COVID (secondary outcome) on day 180 was lower in the metformin group (−0.007; 95% CrI −0.015 to 0.001; PPE 0.96), with a relative risk of 0.495 (95% CrI 0.155 to 0.995).

**Conclusions:** The posterior probability of efficacy for metformin preventing the primary endpoint did not exceed the prespecified threshold of 0.975 for declaring efficacy. Secondary outcomes were numerically better with metformin.

**Trial Registration:** ClinicalTrials.gov (NCT04885530).

## INTRODUCTION

Estimates suggest that approximately 7% of adults in the US have had post-acute sequelae of COVID (PASC), now often referred to as long COVID (LC), making it potentially the sixth most common chronic disease in the US.^1–3^ Waning immunity and the risk of more severe variants continue to pose a risk to the population. Thus, identifying therapies to prevent and treat LC remains important.

The definition of LC has evolved to include either symptom-based or condition-based criteria.^4^ One randomized trial of early outpatient treatment of SARS-CoV-2 enrolled >1000 adults with no prior infection and >50% vaccinated to test whether metformin prevented severe acute COVID-19 or LC compared with placebo.^5^ The trial sent surveys to participants for 10 months after randomization to assess medical provider diagnoses of LC, for which there was a 4.1% absolute reduction of LC for participants randomized to metformin versus placebo.^5^ Confirmatory randomized trials were needed of metformin as a treatment of acute infection to prevent subsequent LC.

The Accelerating Coronavirus Disease 2019 Therapeutic Interventions and Vaccines (ACTIV-6) platform randomized clinical trial evaluated repurposed medications in the outpatient setting.^6^ For this study, the ACTIV-6 platform sought to evaluate the effect of 14 days of immediate-release metformin at the time of acute infection for preventing symptoms attributed to COVID-19 on day 180.

## METHODS

### Trial Design and Oversight

ACTIV-6 was a double-blind, randomized, placebo-controlled platform trial of repurposed medications in outpatient adults with mild to moderate COVID-19.^6,7^ ACTIV-6 was decentralized, allowing virtual enrollment and enrollment through health care and community settings, to improve access. The trial protocol was approved by a central institutional review board with review at each site. Informed consent was obtained from each participant. An independent data and safety monitoring committee oversaw participant safety, efficacy, and trial conduct. Reporting followed CONSORT guidelines for randomized trials.^8^

### Participants

The ACTIV-6 platform was open to enrollment from June 11, 2021 to April 29, 2024, with ∼11,000 participants randomized across 7 interventions. The metformin group enrolled participants from 90 US sites between September 19, 2023 and May 1, 2024 (**Supplemental Appendix**).

Eligibility criteria were confirmed at the site level and included age ≥30 years, confirmed SARS-CoV-2 infection (positive polymerase chain reaction or home-based antigen test) within 10 days, and actively experiencing ≥2 COVID-19 symptoms for ≤7 days at the time of consent. Individuals meeting the following criteria were excluded: current hospitalization for COVID-19; current or recent use (within 14 days) of metformin, insulin, or sulfonylureas; and known allergy or contraindications to metformin. Prior infection, receipt of COVID-19 vaccinations, or current use of approved or emergency use authorization therapeutics for COVID-19 were allowed. The modified intention to treat (mITT) sample in this decentralized trial were participants who remained after excluding those who did not receive delivery of the study drug or were administratively withdrawn (**Figure 1).**

**Figure 1.**
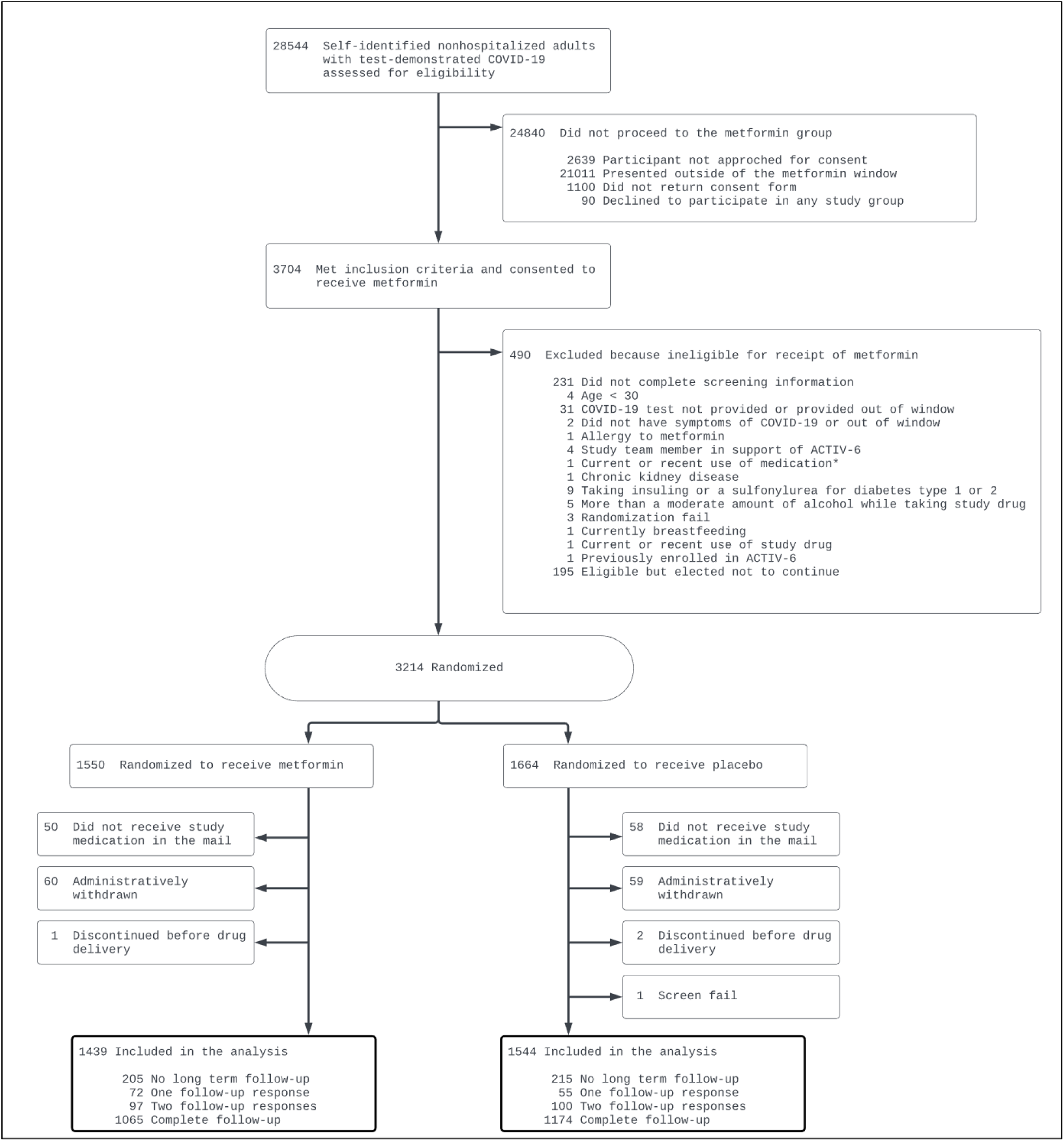
Participant study flow

### Randomization

Enrollment for metformin did not overlap with other agents on the platform. A random number generator completed simple randomization in a 1:1 ratio of metformin or placebo.

### Interventions

Participants received metformin (immediate-release tablets from Aurobindo Pharma USA, Durham, NC) or placebo via home delivery from a centralized pharmacy. A matching placebo became available after November 9, 2023, so 134 participants in the placebo group were enrolled using generic placebo. The 14-day dosing regimen was 500 mg once daily for 1 day; 500 mg twice daily for 4 days; then 500 mg in the morning and 1000 mg in the evening for 9 days (36 tablets total).

### Outcome Measures

The primary outcome was post-acute sequelae of SARS-CoV-2 or death (PASCD), a composite outcome of patient-reported symptoms or mortality, because death is a competing risk for developing LC. For assessing symptoms, participants were asked: “Please choose the response that best describes the severity of your COVID-19 symptoms today,” with options for no symptoms, mild, moderate, and severe. Participants who were deceased were deemed to have experienced the endpoint. Except for participants who reported a new COVID-19 infection within 28 days, participants who reported any COVID-19 symptoms were deemed to have experienced the endpoint.

The primary measure of treatment benefit was the difference in PASCD risk on day 180 between the metformin group and the placebo group, with a difference less than zero indicating a lower risk in the metformin group.

#### Secondary Outcome Measures

The presence of PASCD on days 90 and 120 were secondary outcomes. A participant might meet the criteria for PASCD on follow-up day 90 but not on day 120 or day 180 if they no longer reported symptoms. In addition to summarizing the treatment effect on the absolute scale (as a risk difference), the treatment effect for PASCD was also summarized on the relative scale as a risk ratio.

Symptom burden (none, mild, moderate, severe, deceased) on days 90, 120, or 180 were secondary outcomes. The treatment effect for symptom burden was summarized with common odds ratios.

The number of participants reporting a new infection within 28 days before day 90, 120, or 180, was reported to understand whether symptoms were from a current infection or a past infection (the infection that determined eligibility for the trial or a prior infection).

In addition to outcomes based on symptoms, participants were asked about a clinician diagnosis of LC with the question: “Have you been told by a medical provider that you have long COVID since your last survey?” on days 120 and 180. Participants who responded yes to having received a clinician diagnosis of LC achieved this endpoint, summarized with risk differences and risk ratio.

### Trial Procedures

Current infection status was confirmed by sites. Screening, eligibility, demographic information, medical history, use of concomitant medications, and COVID-19 symptoms were self-reported. The centralized investigational pharmacy packaged and shipped active or placebo study products to participants. Day 1 was defined based on receipt of study product as tracked by the courier. Symptom assessments were sent to participants via the online REDCap electronic data portal on days 90, 120, 150, and 180.

Two safety events of interest were added for the metformin protocol: lactic acidosis or hypoglycemia measured in a medical encounter. Participants were not instructed to monitor blood sugar, and the study did not send glucometers.

### Statistical Analysis Plan

A treatment effect, summarizing the differences between the metformin and placebo groups, was estimated for each outcome. The treatment effect for the primary outcome was the PASCD risk difference on day 180 in the full mITT sample. It was estimated from a Bayesian multivariate model of PASCD at days 90, 120, and 180, based on survey outcomes collected on the same days.

The multivariate model leveraged all available data, incorporating (a) outcome information from every participant who contributed at least 1 long-term follow-up survey and (b) baseline covariate information from all participants. The multivariate model was constructed as a first-order Markov model, comprising 3 covariate-adjusted regression sub-models: (a) day 90 outcomes; (b) day 120 outcomes adjusting for day 90; and (c) day 180 adjusting for day 120. Missing outcome data was accommodated with the observed data likelihood.

Covariate adjustment in each sub-model planned terms for randomization assignment, age as restricted cubic splines, body mass index (BMI) as restricted cubic splines with possible shift at the obesity threshold of 30 kg/m^2^, sex, duration of symptoms before day 1, prior infection, vaccination status, geographic region, calendar date, call center, baseline disease severity. Baseline disease severity was calculated from symptom surveys, using the symptom score captured on or before the day of drug delivery. If the participant received the medication and took it prior to reporting symptoms, their baseline disease severity was the score captured prior to drug delivery. In addition to the planned list of covariates, the plan was updated after the day 28 analysis to include a contingency if fewer than expected outcomes were observed, specifically increasing the degree of regularization via baseline covariate prior distributions, using weakly informative rather than non-informative prior distributions for the baseline covariates, and performing a sensitivity analysis using a reduced number of covariates: obesity, prior COVID-19 infection, and vaccination status (**eTable 1**).

The prior distribution for each model parameter was specified individually. The priors for regression intercept parameters were flat, meaning incidence estimates were unconstrained and estimated from the observed data. All other regression parameters were centered at zero and weakly informative. Computer simulations performed during the planning phase indicated an additional prior on the treatment effect was not needed to achieve type I error control. The posterior distribution was calculated using Markov Chain Monte-Carlo sampling methods and approximated with a multivariate normal distribution for visualizations.

The marginal risk of PASCD on day 180 was derived from the multivariate model for both groups, and the treatment effect was the risk in the metformin group subtracting the risk in the placebo group, marginalizing the difference over the observed covariate distribution. The treatment effect was oriented so that a value less than zero was indicative of a lower incidence of PASCD in the metformin group. The portion of the posterior distribution of the treatment effect less than zero was the posterior probability of efficacy (PPE). The median and highest density interval of the posterior were the point and interval estimates of the treatment effect, respectively.

The same risk differences, calculated at days 90 and 120, were calculated as secondary outcomes. The risk ratio, calculated for days 90, 120, and 180, were also calculated from the same model, as an expression of the treatment effect on the relative scale. Symptom burden odds ratios (OR) at days 90, 120, 180 were also calculated.

Participant-reported clinician diagnosis of LC or death was modeled with the first-order Markov model among participants responding to the day 120 or day 180 surveys, using the same methods for missing outcomes, priors, and calculation of treatment effect estimates. Unadjusted counts, proportions, and ratios of PASCD and clinician diagnosis of LC were tabulated for each follow-up survey. A predefined assessment of treatment effect heterogeneity (HTE) encompassed age, symptom duration, body mass index (BMI), symptom severity on day 1, calendar time (indicative of circulating SARS-CoV-2 variant), sex, and vaccination status.

Because the PPE is inherently a 1-sided measure, the threshold for declaring efficacy was updated after the day 28 analysis to require a PPE of 0.975 for the risk difference of PASCD on day 180. Secondary outcomes were exploratory and not adjusted for multiple comparisons. Analyses were performed with R^10^ version 4.5 with the following primary packages: rstan and rstanarm.^11,129^

## RESULTS

### Study Population

Of the 3214 participants randomized, 231 (7.2%) were administratively withdrawn or excluded because study drug was not delivered within 7 days. Thus, the mITT analysis included 2983 participants, of whom 1439 (48.2%) received metformin and 1544 (51.8%) received placebo (**Figure 1**).

The median age of the mITT population was 47 years (interquartile range [IQR] 38–57); 63.4% were female, 80.1% were White, 11.7% were Black, 0.8% were American Indian or Alaska Native, and 46.5% identified as Hispanic/Latino. Overall, 2% had diabetes, 2% had high blood pressure, 68.3% reported ≥2 doses of a SARS-CoV-2 vaccine, 53.5% reported having at least 1 prior COVID-19 infection, and 38.2% had obesity with 10% more in the metformin than placebo group having obesity (**Table 1**).

**Table 1.** Baseline characteristics.

|  | **Metformin**<br>**(n=1439)** | **Placebo**<br>**(n=1544)** | **Overall**<br>**(n=2983)** |
| --- | --- | --- | --- |
| Age, median (IQR), y | 47 (38-57) | 48.0 (37-58) | 47 (38-58) |
| Age <50 y, No. (%) | 796 (55.3) | 843 (54.6) | 1639 (54.9) |
| Sex, No. (%) |  |  |  |
| Female | 917 (63.7) | 973 (63.02) | 1890 (63.4) |
| Male | 519 (36.1) | 566 (36.66) | 1085 (36.4) |
| Undifferentiated, unknown, prefer not to answer | 3 (0.2) | 5 (0.32) | 8 (0.3) |
| Race,<sup>a</sup> No. (%) |  |  |  |
| American Indian or Alaska Native | 18 (1.3) | 7 (0.45) | 25 (0.8) |
| Asian | 37 (2.6) | 40 (2.59) | 77 (2.6) |
| Black or African American | 164 (11.4) | 184 (11.92) | 348 (11.7) |
| Middle Eastern or North African | 35 (2.4) | 23 (1.49) | 58 (1.9) |
| Native Hawaiian or Other Pacific Islander | 4 (0.3) | 4 (0.26) | 8 (0.3) |
| White | 1158 (80.5) | 1232 (79.79) | 2390 (80.1) |
| None of the above | 31 (2.2) | 37 (2.40) | 68 (2.3) |
| Prefer not to answer | 25 (1.7) | 28 (1.81) | 53 (1.8) |
| Hispanic/Latino, No. (%) | 679 (47.2) | 712 (46.1) | 1391 (46.6) |
| Region, No. (%) |  |  |  |
| Midwest | 295 (20.5) | 308 (20.0) | 603 (20.2) |
| Northeast | 117 (8.1) | 133 (8.6) | 250 (8.4) |
| South | 888 (61.7) | 942 (61.0) | 1830 (61.3) |
| West | 139 (9.7) | 161 (10.43) | 300 (10.1) |
| Call center, No. (%) | 41 (2.8) | 42 (2.72) | 83 (2.8) |
| **Medical history** |  |  |  |
| BMI, median (IQR), kg/m<sup>2</sup> | 28.6 (25.4-32.6) | 28.3 (25.1-31.9) | 28.4 (25.2-32.2) |
| BMI >30 kg/m<sup>2</sup>, No. (%) | 576 (40.0) | 563 (36.5) | 1139 (38.2) |
| Heart disease, No./total (%) | 41/1382 (3.0) | 48/1479 (3.2) | 89/2861 (3.1) |
| Diabetes, No./total (%) | 30/1382 (2.2) | 27/1479 (1.8) | 57/2861 (2.0) |
| High blood pressure, No./total (%) | 281/1382 (20.3) | 316/1479 (21.37) | 597/2861 (20.9) |
| COPD, No./total (%) | 30/1382 (2.2) | 28/1479 (1.9) | 58/2861 (2.0) |
| Asthma, No./total (%) | 141/1382 (10.2) | 135/1479 (9.13) | 276/2861 (9.6) |
| Chronic kidney disease, No./total (%) | 1/1382 (0.1) | 6/1479 (0.4) | 7/2861 (0.2) |
| Smoker, past year, No./total (%) | 196/1382 (14.2) | 229/1479 (15.5) | 425/2861 (14.9) |
| Malignant cancer, No./total (%) | 29/1380 (2.10) | 31/1474 (2.10) | 60/2854 (2.10) |
| Prior infection, No./total (%) | 739/1350 (54.74) | 764/1459 (52.36) | 1503/2809 (53.51) |
| **SARS-CoV-2 vaccine doses, No. (%)** |  |  |  |
| None | 462 (32.1) | 479 (31.0) | 941 (31.5) |
| 1 | 2 (0.14) | 2 (0.13) | 4 (0.13) |
| 2+ | 975 (67.8) | 1063 (68.9) | 2038 (68.3) |
| Any immunity (prior infection or any vaccine), | 1170/1401 (83.5) | 1252/1505 (83.2) | 2422/2906 (83.3) |
| No./total (%) |  |  |  |
| Days between symptom onset and receipt of drug, median (IQR) | 5 (4-7) | 5 (4-7) | 5 (4-7) |
| Days between symptom onset and enrollment, median (IQR) | 3 (2-5) [n=1407] | 3 (2-5) [n=1509] | 3 (2-5) [n=2916] |
| Symptom burden on/before day of drug receipt,<sup>e</sup> No./total (%) |  |  |  |
| None | 67/1405 (4.8) | 71/1508 (4.7) | 138/2913 (4.7) |
| Mild | 624/1405 (44.4) | 675/1508 (44.8) | 1299/2913 (44.6) |
| Moderate | 668/1405 (47.5) | 711/1508 (47.1) | 1379/2913 (47.3) |
| Severe | 46/1405 (3.3) | 51/1508 (3.4) | 97/2913 (3.3) |
| Use of remdesivir, No. (%) | 1 (0.07) | 0 (0.00) | 1 (0.03) |
| Use of nirmatrelvir/ritonavir, No. (%) | 204 (14.2) | 229 (14.8) | 433 (14.5) |
| Use of monoclonal antibodies, No. (%) | 31 (2.2) | 43 (2.8) | 74 (2.5) |
| Use of molnupiravir, No. (%) | 10 (0.7) | 13 (0.8) | 23 (0.8) |

BMI, body mass index (calculated as weight in kilograms divided by height in meters squared); COPD, chronic obstructive pulmonary disease.
<sup>a</sup>Participants were presented with each of these options and may have selected any combination of the race descriptors, including “none of the above” and “prefer not to answer.” Consequently, the sums of counts over all categories do not match the column totals.
<sup>b</sup>The following state groups define each region. Northeast includes Connecticut, Maine, Massachusetts, New Hampshire, Rhode Island, Vermont, New Jersey, New York, and Pennsylvania; Midwest includes Indiana, Illinois, Michigan, Ohio, Wisconsin, Iowa, Kansas, Minnesota, Missouri, Nebraska, North Dakota, and South Dakota; South includes Delaware, District of Columbia, Florida, Georgia, Maryland, North Carolina, South Carolina, Virginia, West Virginia, Alabama, Kentucky, Mississippi, Tennessee, Arkansas, Louisiana, Oklahoma, and Texas; West includes Arizona, Colorado, Idaho, New Mexico, Montana, Utah, Nevada, Wyoming, Alaska, California, Hawaii, Oregon, and Washington.
<sup>c</sup>Patients may have alternatively been recruited at local clinical sites.
<sup>d</sup>Medical history was provided by participants responding to the following prompts: “Has a doctor told you that you have any of the following?” “Have you ever experienced any of the following? (select all that apply),” and “Have you ever smoked tobacco products?”
<sup>e</sup>Each day, participants were asked, “Please choose the response that best describes the severity of your COVID-19 symptoms today.”

The proportion of the mITT population who completed the day 90, 120, and 180 surveys ranged from 79% to 81%. Among those randomized to metformin, 1139 (79%) completed the day 180 survey; among those randomized to placebo, 1251 (81%) completed the day 180 survey (**Table 2, eTables 2–4**).

**Table 2.** Observed counts and percentage points for PASCD and clinician diagnosis of long COVID.

|  |  | Overall<br>(n=2983) | Metformin<br>(n=1439) | Placebo<br>(n=1544) |
| --- | --- | --- | --- | --- |
| Clinician diagnosis of long COVID, n (%) |  |  |  |  |
| Day 120 | Yes | 29 (0.97) | 13 (0.90) | 16 (1.04) |
|  | No | 2387 (80.0) | 1146 (79.6) | 1241 (80.4) |
|  | *missing* | 567 (19.0) | 280 (19.5) | 287 (18.6) |
| Day 180 | Yes | 26 (0.87) | 8 (0.56) | 18 (1.17) |
|  | No | 2362 (79.2) | 1130 (78.5) | 1233 (79.9) |
|  | *missing* | 594 (19.9) | 301 (20.9) | 293 (19.0) |
| PASCD, n (%) |  |  |  |  |
| Day 90 | Yes | 96 (3.2) | 44 (3.1) | 52 (3.4) |
|  | No | 2335 (78.3) | 1119 (77.8) | 1216 (78.8) |
|  | *missing* | 552 (18.5) | 276 (19.2) | 276 (17.9) |
| Day 120 | Yes | 101 (3.4) | 40 (2.8) | 61 (4.0) |
|  | No | 2316 (77.7) | 1119 (77.8) | 1197 (77.5) |
|  | *missing* | 566 (19.0) | 280 (19.5) | 286 (18.5) |
| Day 180 | Yes | 79 (2.7) | 33 (2.3) | 46 (3.0) |
|  | No | 2311 (77.5) | 1106 (76.9) | 1205 (78.0) |
|  | *missing* | 593 (19.9) | 300 (20.9) | 293 (19.0) |

PASCD was the primary endpoint: symptoms attributed to COVID-19 on day 180 or mortality. Of note, there were no deaths.

### Primary Outcome

Overall, 79 (2.6%) participants reported having symptoms that they attributed to COVID-19 on day 180 and there were no observed deaths in the study. Among participants randomized to metformin, 33 (2.3%) reported symptoms on day 180; among participants randomized to placebo, 46 (3.0%) reported symptoms on day 180. The covariate-adjusted risk of PASCD was −0.008 (95% credible interval [CrI] −0.022 to 0.006; PPE: 0.83), lower in the metformin group than the placebo group (**Tables 2 & 3**).

### Secondary Outcomes

Overall, 96 (3.2%) participants reported COVID-19 symptoms on day 90, and 101 (3.4%) on day 120. On day 90, 44 (3.1%) participants in the metformin group and 51 (3.4%) in the placebo group reported COVID-19 symptoms; and on day 120, 40 (2.8%) in the metformin group and 61 (4.0%) in the placebo group reported COVID-19 symptoms (**Table 3**).

**Table 3.** Model-based estimate of the treatment effect on the risk difference scale for the primary and secondary outcomes.

| Outcome | Metformin<br>(n=1439) | Placebo<br>(n=1544) | Treatment Effect Estimate <sup>a</sup><br>$\Delta$ (95% CrI)<sup>b</sup> | Posterior<br>P(efficacy) |
| --- | --- | --- | --- | --- |
| **Primary endpoint: PASCD** <sup>c,f</sup> |  |  |  |  |
| Day 180 | 0.029 (0.020, 0.039) | 0.036 (0.027, 0.047) | -0.008 (-0.022, 0.006) | 0.83 |
| *Secondary time points* |  |  |  |  |
| Day 90 | 0.039 (0.029, 0.050) | 0.043 (0.033, 0.055) | -0.003 (-0.019, 0.012) | 0.66 |
| Day 120 | 0.036 (0.025, 0.046) | 0.047 (0.037, 0.059) | -0.011 (-0.027, 0.004) | 0.93 |
| **PASCD** <sup>d,f</sup> |  |  |  |  |
| Day 90 | 0.039 (0.028, 0.050) | 0.041 (0.031, 0.053) | -0.003 (-0.019, 0.013) | 0.63 |
| Day 120 | 0.035 (0.025, 0.046) | 0.048 (0.037, 0.060) | -0.012 (-0.028, 0.004) | 0.94 |
| Day 180 | 0.029 (0.020, 0.039) | 0.036 (0.026, 0.046) | -0.007 (-0.021, 0.006) | 0.85 |
| **Secondary endpoints:** |  |  |  |  |
| **Clinician Diagnosis of long COVID** <sup>d,f</sup> |  |  |  |  |
| Day 120 | 0.011 (0.006, 0.017) | 0.012 (0.007, 0.019) | -0.001 (-0.010, 0.007) | 0.63 |
| Day 180 | 0.007 (0.003, 0.012) | 0.014 (0.008, 0.021) | -0.007 (-0.015, 0.001) | 0.96 |
| **Symptom burden, relative risk scale** <sup>d</sup> |  |  | **Odds Ratio (95% CrI)** <sup>e</sup> |  |
| Day 90 (n=2431) |  |  | 0.90 (0.57, 1.34) | 0.69 |
| Day 120 (n=2417) |  |  | 0.65 (0.43, 0.95) | 0.98 |
| Day 180 (n=2390) |  |  | 0.86 (0.51, 1.32) | 0.74 |

Abbreviations: $\Delta$ , Risk difference; CrI, credible interval; P, probability; PASCD, the primary endpoint was symptoms attributed to COVID-19 on Day 180 or mortality. Of note, no deaths occurred.
<sup>a</sup> Estimates derived from Bayesians model. Prior distributions for intercept terms were flat and non-informative. Prior distributions for covariate terms were weakly informative when included in the model. All outcomes are participant-reported.
<sup>b</sup> Unless otherwise noted, a highest-density credible interval is shown. $\Delta$ that is less than 0 is in the direction of benefit for metformin.
<sup>c</sup> Adjusted for covariates: age, body mass index, calendar date, gender, prior infection, duration of symptoms prior to treatment, vaccination status, geographic region (Northeast, Midwest, South, and West), baseline symptom burden, and an indicator for enrollment to a remote site using baseline characteristics for the full modified intention to treat cohort.
<sup>d</sup> Not covariate adjusted.
<sup>e</sup> An odds ratio < 1.0 favors metformin.
<sup>f</sup> Missing responses were handled using an observed data likelihood.

The covariate-adjusted risk of PASCD on day 90 was −0.003 (95% CrI −0.019 to 0.012; PPE 0.66), lower in the metformin group; and on day 120, the risk was −0.011 (95% CrI −0.027 to 0.004; PPE: 0.93) (**Table 3**).

Overall, on day 120, 29 (0.97%) participants reported having received a clinician diagnosis of LC, 13 (0.90%) in the metformin group and 16 (1.04%) in the placebo group; the risk was −0.001 (−0.010 to 0.007; PPE: 0.63), lower in the metformin group (risk ratio [RR] 0.879; 95% CrI 0.349 to 1.632) (**Table 3**, **Figure 2**). On Day 180, 26 (0.87%) participants reported receiving a clinician diagnosis of LC, 8 (0.56%) in the metformin group and 18 (1.17%) in the placebo group, the risk was −0.007 (95% CrI −0.015 to 0.001; PPE 0.96), RR 0.50 (95% CrI 0.16 to 0.99) (**Table 3**, **Figure 2**).

**Figure 2.**
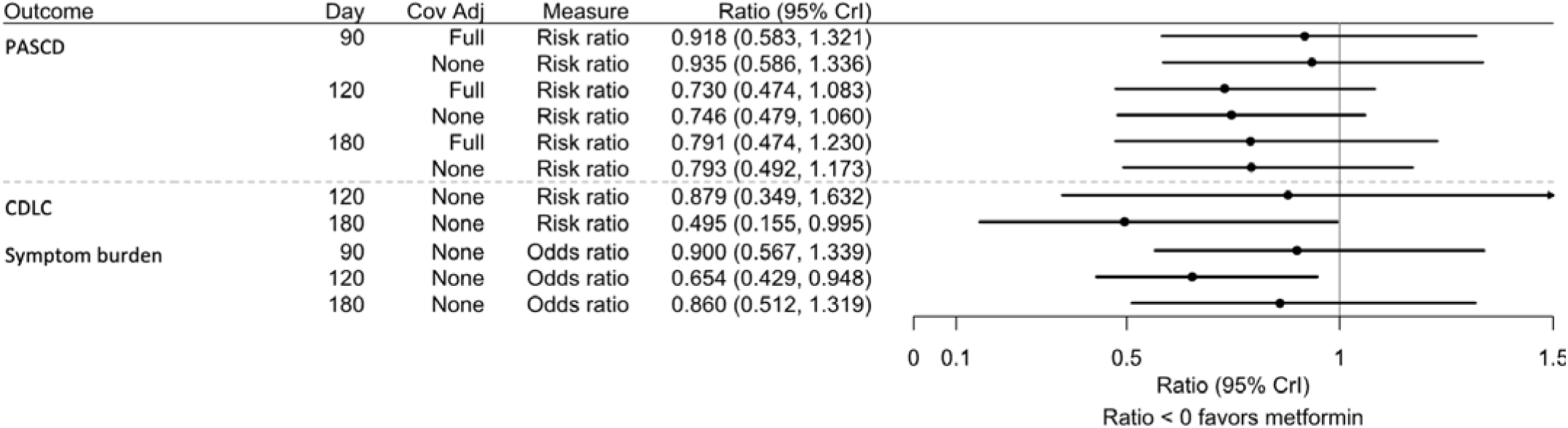
Relative risks for the primary and secondary outcomes The rows for PASCD and clinician diagnosis of Long COVID display the relative risk calculated from Bayesian models of the respective longitudinal outcomes using all available outcome and covariate data. Missing outcome data was handled with the complete data likelihood. Prior distributions for the intercept were non-informative and were weakly informative for all other parameters. Full covariate adjustment included the following covariates: age, body mass index, calendar date, gender, prior infection, duration of symptoms prior to treatment, vaccination status, geographic region (Northeast, Midwest, South, and West), baseline symptom burden, and an indicator for enrollment to a remote site), from the full modified intention to treat sample. The rows for symptom burden report the odds ratios at each time point, calculated from Bayesian proportional odds ordinal outcome models using the subset of participants who responded to the survey for the day in question. Symptom burden is an ordinal outcome with none, mild, moderate, and severe as possible responses. An odds ratio less than one indicates a treatment benefit for metformin. All outcomes are participant-reported. *Abbreviations:* PASCD, the primary endpoint was symptoms attributed to COVID-19 on Day 180 or mortality, of note there were no deaths observed; CDLC=clinician diagnosed Long COVID; CrI, credible interval.

There was no evidence of heterogeneity of treatment effect (**eFigure 1, eTables 5–6**). Posterior distributions are presented in **eFigures 2–7**. For the two safety events of interest for this appendix, there were no episodes of lactic acidosis and there were 6 episodes of participant-reported hypoglycemia reported by sites, 2 in the metformin group and 4 in the placebo group.

## DISCUSSION

Among low-risk outpatient adults with mild to moderate COVID-19, the posterior probability that treatment with immediate-release metformin would prevent symptoms participants attributed to COVID-19 on day 180 was 0.81, which did not meet the predefined 0.975 threshold for efficacy. Secondary outcomes were numerically better with metformin, including a PPE of 0.98 for symptom burden on day 120 and 0.96 for clinician diagnosis of LC on day 180.

In the context of other literature, the direction of these results is consistent with existing data on the use of metformin at the time of acute infection and subsequent development of PASCD. For the death component of PASCD, meta-analyses in adults with type 2 diabetes reported that prevalent metformin use was associated with an approximately 35% lower risk of mortality.^10,11^ A retrospective analysis of medication administered to nursing home residents, verified by bar-code, in the Veterans Health Administration (VHA) reported a hazard ratio (HR) for mortality of 0.48 (95% confidence interval [CI] 0.28 to 0.84) in those taking metformin.^12^ Another VHA analysis reported a relative risk of 0.33 (95% CI 0.25 to 0.43) for metformin use, similar to a prospective cohort in adults with type 2 diabetes.^13,14^ These studies reflect an earlier time in the pandemic when mortality was higher; death is an uncommon outcome in the ACTIV-6 population.

For the LC component of PASCD, observational studies of prevalent metformin use in adults with type 2 diabetes have reported a HR of 0.72 (95% CI 0.67 to 0.77) when LC was ascertained using natural language processing from the electronic health record; a HR of 0.79 (95% CI 0.71 to 0.88) when LC was ascertained using the LC diagnosis code in the electronic health record; and an odds ratio of 0.32 (0.13 to 0.73) when LC was prospectively ascertained with participant reported surveys.^15–17^

In adults without type 2 diabetes, a target trial emulation study reported a HR of 0.47 (95% CI 0.25 to 0.89) for new use of metformin within a week of infection and developing LC, defined using diagnosis code or computable phenotype.^18^ In the COVID-OUT randomized trial, the HR for participant-reported clinician diagnosis of LC in those randomized to 14 days of metformin was 0.59 (95% CI 0.39 to 0.89), and medical records were obtained to confirm these participant-reported diagnoses.^5^

In ACTIV-6, the presence of symptoms and incidence of clinician diagnoses of LC on day 180 was lower than these observational and prospective studies, likely indicating a favorable evolution of COVID-19 severity. A lower incidence of LC may be due to prior immunity, as ACTIV-6 is unique in that persons with prior infection were eligible. Thus, over 83% of participants had some prior immunity: 54% reported having had at least 1 prior infection and 68% had received at least 2 doses of a SARS-CoV-2 vaccine. Additionally, most participants were enrolled when the JN-1 variant dominated, which was a variant with higher infectivity but lower morbidity than previous and subsequent variants.^19–23^ The lower overall incidence of symptoms may also be due to the ACTIV-6 survey only capturing symptoms on the day of the survey because symptoms can be intermittent.

A consistent mechanistic finding is that SARS-CoV-2 activates the mammalian target of rapamycin complex 1 (mTORC1), which decreases the epithelial barrier function in the gut.^24–26^ Dysfunctional gut epithelium has been implicated in LC via dysregulated signaling to the brain.^27,28^ The virus also decreases short-chain fatty acid (SCFA)-producing bacteria, which has strongly correlated with LC at 6 months.^29–32^ These changes in the gut can leak harmful proteins that cause disease over time.^33^ This could explain why some develop LC after a symptom-free period and potentially explain why the total number of participants reporting symptoms was higher at day 120 than day 90 in this trial. Metformin is known to inhibit mTOR in the gut and increase the relative abundance of SCFA-producing bacteria, actions that might be protective during acute and long-term morbidity.^34–36^

This is the first phase 3 trial to randomize adults without chronic disease or disease risk to metformin. There were fewer reported episodes of low blood sugar in the metformin group and no reported lactic acidosis during the full follow-up period.

### Limitations

This study does have limitations. It is possible that the COVID-19 symptoms that participants reported on day 90, 120, or 180 were LC symptoms that preceded their enrollment in ACTIV-6. The proportion of missing data for the primary and secondary outcomes was around 20% for the day 90, 120, and 180 surveys. This study had a higher proportion of post-randomization exclusions compared with earlier studies, and the placebo group is more than 7% larger than the metformin group. Metformin has antiviral actions against SARS-CoV-2, enrollment of participants later in the course of infection than trials of other SARS-CoV-2 antivirals may have impacted the results.^37–45^

### Conclusions

Among low-risk outpatient adults with SARS-CoV-2 infection and a high degree of prior immunity, the posterior probability of efficacy for metformin preventing the primary endpoint did not exceed the prespecified threshold for declaring efficacy. This sample was enrolled during a variant that was milder that previous and subsequent variants of SARS-CoV-2, and the proportion of participants with COVID-19 symptoms or clinician diagnosis of LC at 6 months was low. The posterior probability of efficacy for metformin was high for preventing a clinician diagnosis of LC and for symptom burden, consistent with prior clinical trial and observational data. There were no safety concerns in this trial of metformin in low-risk adults.

## Data Availability

ACTIV-6 is a platform trial using shared placebos. On completion of the platform trial, when there is no risk of unblinding across study arms, the data will be made publicly available by depositing it in an approved data repository such as NHLBI BioData Catalyst.

## Acknowledgments

We thank George J. Hanna, MD, of the Biomedical Advanced Research and Development Authority, for his roles in trial implementation and operations. We also thank the ACTIV-6 Data Monitoring Committee, Clinical Events Committee, and Stakeholder Advisory Committee Members (listed below) for their contributions.

Data Monitoring Committee: Clyde Yancy, MD, MSc, Northwestern University Feinberg School of Medicine; Adaora Adimora, MD, University of North Carolina, Chapel Hill; Susan Ellenberg, PhD,

University of Pennsylvania; Kaleab Abebe, PhD, University of Pittsburgh; Arthur Kim, MD, Massachusetts General Hospital; John D. Lantos, MD, Children’s Mercy Hospital; Jennifer Silvey-Cason, Participant representative; Frank Rockhold, PhD, Duke Clinical Research Institute; Sean O’Brien, PhD, Duke Clinical Research Institute; Frank Harrell, PhD, Vanderbilt University Medical Center; Zhen Huang, MS, Duke Clinical Research Institute.

Clinical Events Committee: Renato Lopes, MD, PhD, MHS, W. Schuyler Jones, MD, Antonio Gutierrez, MD, Robert Harrison, MD, David Kong, MD, Robert McGarrah, MD, Michelle Kelsey, MD, Konstantin Krychtiuk, MD, Vishal Rao, MD all of the Duke Clinical Research Institute, Duke University School of Medicine.

Stakeholder Advisory Committee: Megan E. Hamm, Kathleen McTigue, Kirk T. Phillips, Andrew Vasey, Talethia Edwards, Danielle Nelson, Greg Merritt, Jeannie Nguyen, Josh Denson, Jonathan Arnold, Matthew W. McCarthy, Florence Thicklin.

Elizabeth E.S. Cook of the Duke Clinical Research Institute provided editorial support.

## Author Contributions

Drs Naggie, Hernandez, and Lindsell had full access to all the blinded data in the study. Dr Stewart was provided curated study data and takes responsibility for the integrity of the data analysis. All authors contributed to the drafting and review of the manuscript and agreed to submit for publication.

## References

1. Fang Z, Ahrnsbrak R, Rekito A. Evidence Mounts That About 7% of US Adults Have Had Long COVID. JAMA. 2024;332(1):5–6. doi:10.1001/jama.2024.11370

2. Geng LN, Bonilla H, Hedlin H, et al. Nirmatrelvir-Ritonavir and Symptoms in Adults With Postacute Sequelae of SARS-CoV-2 Infection: The STOP-PASC Randomized Clinical Trial. JAMA Intern Med. 2024;184(9):1024–1034. doi:10.1001/jamainternmed.2024.2007

3. Telesford I, McGough M, Tevis D, Cotter L. How has the burden of chronic diseases in the U.S. and peer nations changed over time? https://www.healthsystemtracker.org/chart-collection/how-has-the-burden-of-chronic-diseases-in-the-u-s-and-peer-nations-changed-over-time/#Age-standardized%20percent%20of%20adults%20with%20obesity,%201990-2022

4. Ely EW, Brown Lisa M, Fineberg Harvey V. Long Covid Defined. N Engl J Med. 2024/11/06 2024;391(18):1746–1753. doi:10.1056/NEJMsb2408466

5. Bramante CT, Buse JB, Liebovitz DM, et al. Outpatient treatment of COVID-19 and incidence of post-COVID-19 condition over 10 months (COVID-OUT): a multicentre, randomised, quadruple-blind, parallel-group, phase 3 trial. Lancet Infect Dis. Jun 8 2023;doi:10.1016/S1473-3099(23)00299-2

6. ACTIV-6: Operationalizing a decentralized, outpatient randomized platform trial to evaluate efficacy of repurposed medicines for COVID-19. J Clin Transl Sci. 2023;7(1):e221. doi:10.1017/cts.2023.644

7. Naggie S, Boulware DR, Lindsell CJ, et al. Effect of Ivermectin vs Placebo on Time to Sustained Recovery in Outpatients With Mild to Moderate COVID-19: A Randomized Clinical Trial. JAMA. 2022;328(16):1595–1603. doi:10.1001/jama.2022.18590

8. Schulz KF, Altman DG, Moher D, Group C. CONSORT 2010 Statement: updated guidelines for reporting parallel group randomised trials. Trials. Mar 24 2010;11:32. doi:10.1186/1745-6215-11-32

9. Team SD. RStan: the R interface to Stan. R package version 2.32.7. https://mc-stan.org/

10. Song ZH, Huang QM, Xu SS, Zhou JB, Zhang C. The Effect of Antihyperglycemic Medications on COVID-19: A Meta-analysis and Systematic Review from Observational Studies. Ther Innov Regul Sci. Jul 2024;58(4):773–787. doi:10.1007/s43441-024-00633-6

11. Lukito AA, Pranata R, Henrina J, Lim MA, Lawrensia S, Suastika K. The Effect of Metformin Consumption on Mortality in Hospitalized COVID-19 patients: a systematic review and meta-analysis. Diabetes Metab Syndr. Nov-Dec 2020;14(6):2177–2183. doi:10.1016/j.dsx.2020.11.006

12. Lally MA, Tsoukas P, Halladay CW, O’Neill E, Gravenstein S, Rudolph JL. Metformin is Associated with Decreased 30-Day Mortality Among Nursing Home Residents Infected with SARS-CoV2. J Am Med Dir Assoc. Jan 2021;22(1):193–198. doi:10.1016/j.jamda.2020.10.031

13. Hunt CM, Efird JT, Redding TS, et al. Medications Associated with Lower Mortality in a SARS-CoV-2 Positive Cohort of 26,508 Veterans. J Gen Intern Med. 2022/12/01 2022;37(16):4144–4152. doi:10.1007/s11606-022-07701-3

14. Usman A, Bliden KP, Cho A, et al. Metformin use in patients hospitalized with COVID-19: lower inflammation, oxidative stress, and thrombotic risk markers and better clinical outcomes. J Thromb Thrombolysis. Feb 2022;53(2):363–371. doi:10.1007/s11239-022-02631-7

15. Soff S, Yoo YJ, Bramante C, et al. Association of glycemic control with Long COVID in patients with type 2 diabetes: findings from the National COVID Cohort Collaborative (N3C). BMJ Open Diabetes Res Care. Feb 4 2025;13(1)doi:10.1136/bmjdrc-2024-004536

16. Johnson SGW, M.; Sturmer, T, et al. Prevalent Metformin Use in Adults with Diabetes and the Incidence of Long Covid: An EHR-based Cohort Study from the RECOVER Program. In preparation. 2024;

17. Mateu L, Tebe C, Loste C, et al. Determinants of the onset and prognosis of the post-COVID-19 condition: a 2-year prospective observational cohort study. Lancet Reg Health Eur. Oct 2023;33:100724. doi:10.1016/j.lanepe.2023.100724

18. Bramante CT. Metformin reduces the risk of Long COVID or Death over 6 months in an Emulated Target Trial of Primarily Omicron-infected Adults without Diabetes or Prediabetes: a New-User, Active-Comparator Analysis Using the National COVID Cohort Collaborative (N3C) Electronic Health Record Database. 2024:

19. Ma KC, Castro J, Lambrou AS, et al. Genomic Surveillance for SARS-CoV-2 Variants: Circulation of Omicron XBB and JN.1 Lineages - United States, May 2023-September 2024. MMWR Morb Mortal Wkly Rep. Oct 24 2024;73(42):938–945. doi:10.15585/mmwr.mm7342a1

20. Lewnard JA, Mahale P, Malden D, et al. Immune escape and attenuated severity associated with the SARS-CoV-2 BA.2.86/JN.1 lineage. Nat Commun. Oct 3 2024;15(1):8550. doi:10.1038/s41467-024-52668-w

21. Levy ME, Chilunda V, Davis RE, et al. Reduced Likelihood of Hospitalization with the JN.1 or HV.1 SARS-CoV-2 Variants Compared to the EG.5 Variant. J Infect Dis. Jul 19 2024;doi:10.1093/infdis/jiae364

22. Qu P, Evans JP, Faraone JN, et al. Enhanced neutralization resistance of SARS-CoV-2 Omicron subvariants BQ.1, BQ.1.1, BA.4.6, BF.7, and BA.2.75.2. Cell Host Microbe. Jan 11 2023;31(1):9–17 e3. doi:10.1016/j.chom.2022.11.012

23. Wang Q, Mellis IA, Ho J, et al. Recurrent SARS-CoV-2 spike mutations confer growth advantages to select JN.1 sublineages. Emerg Microbes Infect. Dec 2024;13(1):2402880. doi:10.1080/22221751.2024.2402880

24. Zambalde É P, Dias TL, Maktura GC, et al. Increased mTOR Signaling and Impaired Autophagic Flux Are Hallmarks of SARS-CoV-2 Infection. Curr Issues Mol Biol. Dec 31 2022;45(1):327–336. doi:10.3390/cimb45010023

25. Xie Y, Zhao Y, Shi L, et al. Gut epithelial TSC1/mTOR controls RIPK3-dependent necroptosis in intestinal inflammation and cancer. J Clin Invest. 04/01/ 2020;130(4):2111–2128. doi:10.1172/JCI133264

26. Gutiérrez-Martínez IZ, Rubio JF, Piedra-Quintero ZL, et al. mTORC1 Prevents Epithelial Damage During Inflammation and Inhibits Colitis-Associated Colorectal Cancer Development. Transl Oncol. Jan 2019;12(1):24–35. doi:10.1016/j.tranon.2018.08.016

27. Wong AC, Devason AS, Umana IC, et al. Serotonin reduction in post-acute sequelae of viral infection. Cell. Oct 26 2023;186(22):4851–4867.e20. doi:10.1016/j.cell.2023.09.013

28. Giron LB, Peluso MJ, Ding J, et al. Markers of fungal translocation are elevated during post-acute sequelae of SARS-CoV-2 and induce NF-κB signaling. JCI Insight. Aug 8 2022;7(15)doi:10.1172/jci.insight.160989

29. Zhang F, Lau RI, Liu Q, Su Q, Chan FKL, Ng SC. Gut microbiota in COVID-19: key microbial changes, potential mechanisms and clinical applications. Nat Rev Gastroenterol Hepatol. May 2023;20(5):323–337. doi:10.1038/s41575-022-00698-4

30. Hamrefors V, Kahn F, Holmqvist M, et al. Gut microbiota composition is altered in postural orthostatic tachycardia syndrome and post-acute COVID-19 syndrome. Sci Rep. 2024/02/09 2024;14(1):3389. doi:10.1038/s41598-024-53784-9

31. Basting CM, Langat R, Broedlow CA, et al. SARS-CoV-2 infection is associated with intestinal permeability, systemic inflammation, and microbial dysbiosis in hospitalized COVID-19 patients. bioRxiv. 2023:2023.12.07.570670. doi:10.1101/2023.12.07.570670

32. Liu Q, Mak JWY, Su Q, et al. Gut microbiota dynamics in a prospective cohort of patients with post-acute COVID-19 syndrome. Gut. 2022;71(3):544–552. doi:10.1136/gutjnl-2021-325989

33. DeBari MK, Abbott RD. Adipose Tissue Fibrosis: Mechanisms, Models, and Importance. Int J Mol Sci. Aug 21 2020;21(17)doi:10.3390/ijms21176030

34. Nistala R, Raja A, Pulakat L. mTORC1 inhibitors rapamycin and metformin affect cardiovascular markers differentially in ZDF rats. Can J Physiol Pharmacol. 2017;95(3):281–287. doi:10.1139/cjpp-2016-0567%M28177677

35. Planas D, Pagliuzza A, Ponte R, et al. LILAC pilot study: Effects of metformin on mTOR activation and HIV reservoir persistence during antiretroviral therapy. EBioMedicine. Mar 2021;65:103270. doi:10.1016/j.ebiom.2021.103270

36. Placeholder. CB will try to get the COVID-OUT stool samples analyzed.

37. Jayk Bernal A, Gomes da Silva MM, Musungaie DB, et al. Molnupiravir for Oral Treatment of Covid-19 in Nonhospitalized Patients. N Engl J Med. Feb 10 2022;386(6):509–520. doi:10.1056/NEJMoa2116044

38. Hammond J, Leister-Tebbe H, Gardner A, et al. Oral Nirmatrelvir for High-Risk, Nonhospitalized Adults with Covid-19. N Engl J Med. 2022;386(15):1397–1408. doi:10.1056/NEJMoa2118542

39. Cao B, Wang Y, Lu H, et al. Oral Simnotrelvir for Adult Patients with Mild-to-Moderate Covid-19. N Engl J Med. 2024/01/17 2024;390(3):230–241. doi:10.1056/NEJMoa2301425

40. Gordon DE, Jang GM, Bouhaddou M, et al. A SARS-CoV-2 protein interaction map reveals targets for drug repurposing. Nature. 2020/07/01 2020;583(7816):459–468. doi:10.1038/s41586-020-2286-9

41. Parthasarathy H, Tandel D, Siddiqui AH, Harshan KH. Metformin suppresses SARS-CoV-2 in cell culture. Virus Res. Nov 20 2022;323:199010. doi:10.1016/j.virusres.2022.199010

42. Ventura-López C, Cervantes-Luevano K, Aguirre-Sánchez JS, et al. Treatment with metformin glycinate reduces SARS-CoV-2 viral load: An in vitro model and randomized, double-blind, Phase IIb clinical trial. Biomed Pharmacother. Aug 2022;152:113223. doi:10.1016/j.biopha.2022.113223

43. Bramante CT, Beckman KB, Mehta T, et al. Favorable Antiviral Effect of Metformin on Severe Acute Respiratory Syndrome Coronavirus 2 Viral Load in a Randomized, Placebo-Controlled Clinical Trial of Coronavirus Disease 2019. Clin Infect Dis. May 1 2024;doi:10.1093/cid/ciae159

44. Schaller MA, Sharma Y, Dupee Z, et al. Ex vivo SARS-CoV-2 infection of human lung reveals heterogeneous host defense and therapeutic responses. JCI Insight. 09/22/ 2021;6(18)doi:10.1172/jci.insight.148003

45. Xian H, Liu Y, Rundberg Nilsson A, et al. Metformin inhibition of mitochondrial ATP and DNA synthesis abrogates NLRP3 inflammasome activation and pulmonary inflammation. Immunity. Jul 13 2021;54(7):1463–1477.e11. doi:10.1016/j.immuni.2021.05.004

