## Supplemental Appendix for "Metformin on the Presence of COVID-19 Symptoms Over 6 Months: The ACTIV-6 Randomized Clinical Trial"

**ACTIV-6 Executive Committee**

G. Michael Felker, Duke Clinical Research Institute (Medical Monitor, blinded)

Allison DeLong, Duke Clinical Research Institute

Rhonda Wilder, Duke Clinical Research Institute

Christopher J. Lindsell, Duke Clinical Research Institute (Data Coordinating Center Executive Director)

George Hanna, Biomedical Advanced Research and Development Authority

**ACTIV-6 Protocol Oversight Committee**

David Boulware, University of Minnesota, POC Co-Chair

Carolyn Bramante, University of Minnesota, POC Co-Chair for the appendix.

Adrian F. Hernandez, Duke Clinical Research Institute (Clinical Coordinating Center PI)

Susanna Naggie, Duke Clinical Research Institute (Clinical Coordinating Center co-PI)

G. Michael Felker, Duke Clinical Research Institute (Medical Monitor, blinded)

Allison DeLong, Duke Clinical Research Institute

Rhonda Wilder, Duke Clinical Research Institute

Christopher J. Lindsell, Duke Clinical Research Institute (Data Coordinating Center Executive Director)

Russell L. Rothman, Vanderbilt University Medical Center (Data Coordinating Center PI)

Thomas G. Stewart, University of Virginia

Sean Collins, Vanderbilt University Medical Center

Sarah Dunsmore, National Center for Advancing Translational Sciences

Stacey Adam, Foundation for the National Institutes of Health

Florence Thicklin, Stakeholder Advisory Committee

Matthew William McCarthy, Weill Cornell Medicine, Stakeholder Advisory Committee

George Hanna, Biomedical Advanced Research and Development Authority

Adit Ginde, University of Colorado Denver – Anschutz

Mario Castro, University of Kansas Medical Center

Dushyantha Jayaweera, University of Miami

Mark Sulkowski, John Hopkins University

Nina Gentile, Lewis Katz School of Medicine at Temple University

Kathleen McTigue, University of Pittsburgh Medical Center

Kim Marschhauser, PCORI

**ACTIV-6 Clinical Trial Team**

Adrian F. Hernandez, Duke Clinical Research Institute (Clinical Coordinating Center PI)

Susanna Naggie, Duke Clinical Research Institute (Clinical Coordinating Center Co-PI)

G. Michael Felker, Duke Clinical Research Institute (Medical Monitor, blinded)

Allison DeLong, Duke Clinical Research Institute

Rhonda Wilder, Duke Clinical Research Institute

Christopher J. Lindsell, Duke Clinical Research Institute (Data Coordinating Center Executive Director)

Sean Collins, Vanderbilt University Medical Center (Data Coordinating Center PI)

Thomas G. Stewart, University of Virginia

**ACTIV-6 Clinical Coordinating Center**

Adrian F. Hernandez, Duke Clinical Research Institute (Clinical Coordinating Center PI)

Susanna Naggie, Duke Clinical Research Institute (Clinical Coordinating Center Co-PI)

G. Michael Felker, Duke Clinical Research Institute (Medical Monitor, blinded)

Allison DeLong, Duke Clinical Research Institute

Rhonda Wilder, Duke Clinical Research Institute

Ryan Fraser, Duke Clinical Research Institute

Mark Ward, Duke Clinical Research Institute

Sarah Weaver, Duke Clinical Research Institute

M. Patricia McAdams, Duke Clinical Research Institute

Kayla Korzekwinski, Duke Clinical Research Institute

Martina Oyelakin, Duke Clinical Research Institute

Samantha Dockery, Duke Clinical Research Institute

Rodney Adkins, Duke Clinical Research Institute

Mathew Crow, Duke Clinical Research Institute

Shanee Light, Duke Clinical Research Institute

Renee Morris, Duke Clinical Research Institute

Erin Nowell, Duke Clinical Research Institute

Kadie Wells, Duke Clinical Research Institute

Alicia Herbert, Duke Clinical Research Institute

Allegra Stone, Duke Clinical Research Institute

Heather Heavlin, Duke Clinical Research Institute

Linley Brown, Duke Clinical Research Institute

Shelley Brunson, Duke Clinical Research Institute

Tina Harding, Duke Clinical Research Institute

Amanda Harrington, Duke Clinical Research Institute

Meaghan Beauchaine, Duke Clinical Research Institute

Kelly Lindblom, Duke Clinical Research Institute

Andrea Burns, Duke Clinical Research Institute

Ahmad Mourad, Duke Clinical Research Institute

**ACTIV-6 Stakeholder Advisory Committee**

Megan E. Hamm

Kathleen McTigue

Kirk T. Phillips

Andrew Vasey

Talethia Edwards

Danielle Nelson

Greg Merritt

Jeannie Nguyen

Josh Denson

Jonathan Arnold

Matthew McCarthy

Florence Thicklin

**ACTIV-6 Independent Data Monitoring Committee**

**Voting Members:**

Clyde Yancy, Northwestern University Feinberg School of Medicine (Chair)

Adaora Adimora, University of North Carolina, Chapel Hill (Vice-chair)

Susan Ellenberg, University of Pennsylvania

Kaleab Abebe, University of Pittsburgh

Arthur Kim, Massachusetts General Hospital

John D. Lantos, Children’s Mercy Hospital

Jennifer Silvey-Cason, Participant representative

**Statistical Data Analysis Center:**

Frank Rockhold, Duke Clinical Research Institute (Lead faculty statistician)

Sean O’Brien, Duke Clinical Research Institute (Faculty statistician)

Frank Harrell, Vanderbilt University Medical Center (DCC-SDAC Faculty Liaison)

Zhen Huang, Duke Clinical Research Institute (Lead statistician)

Hayley Nemeth, Duke Clinical Research Institute

**ACTIV-6 Clinical Events Classification Committee**

Renato Lopes, (CEC PI)

**Faculty Reviewers:** W. Schuyler Jones, Antonio Gutierrez, Robert Harrison, David Kong, Robert McGarrah, Michelle Kelsey, Brad Kolls, Cina Sasannejad, Rajendra (Raj) Mehta,

**Fellow Reviewers:** Mark Kittipibul

**ACTIV-6 Data Coordinating Center**

David Aamodt, Vanderbilt University Medical Center

Deborah Clark, Vanderbilt University Medical Center

Jess Collins, Vanderbilt University Medical Center

Sean Collins, Vanderbilt University Medical Center (Data Coordinating Center PI)

Sheri Dixon, Vanderbilt University Medical Center

Yue Gao, Vanderbilt University Medical Center

John Graves, Vanderbilt University Medical Center

James Grindstaff, Vanderbilt University Medical Center

Frank Harrell, Vanderbilt University Medical Center (DCC-SDAC Faculty Liaison)

Jessica Lai, Vanderbilt University Medical Center

Vicky Liao, Vanderbilt University Medical Center

Christopher J. Lindsell, Duke Clinical Research Institute (Data Coordinating Center Executive Director)

Itzel Lopez, Vanderbilt University Medical Center

Elizabeth Manis, Vanderbilt University Medical Center

Kalley Mankowski, Vanderbilt University Medical Center

Jessica Marlin, Vanderbilt University Medical Center

Alyssa Merkel, Vanderbilt University Medical Center

Sam Nwosu, Vanderbilt University Medical Center

Savannah Obregon, Vanderbilt University Medical Center

Dirk Orozco, Vanderbilt University Medical Center

Nelson Prato, Vanderbilt University Medical Center

Max Rohde, Vanderbilt University Medical Center

Russell Rothman, Vanderbilt University Medical Center

Jana Shirey-Rice, Vanderbilt University Medical Center

Krista Vermillion, Vanderbilt University Medical Center

Jacob Smith, Vanderbilt University Medical Center

Thomas Stewart, Vanderbilt University Medical Center / University of Virginia (Lead statistician)

Hsi-nien Tan, Vanderbilt University Medical Center

Meghan Vance, Vanderbilt University Medical Center

Maria Weir, Vanderbilt University Medical Center

**ACTIV-6 Site Investigators & Study Coordinators**

**Advanced Medical Care, Ltd:** Ray Bianchi, Jen Premas

**Ananda Medical Clinic:** Madhu Gupta, Greg Karawan, Santia Lima, Carey Ziomek

**Arena Medical Group:** Joseph Arena, Sonaly DeAlmeida

**Ascension St. John** Anuj Malik, Jane Bryce, Sarah Swint

**Chandler Regional Medical Center:** Brian Tiffany, Charlotte Tanner, Allegra Sahelian

**Christ the King Health Care, P.C.:** Constance George-Adebayo, Adeolu Adebayo

**Christus St. Vincent Regional Medical Center:** Theresa Ronan, Ashley Woods, Christopher Gallegos, Tamara Flys, Olivia Sloan

**Clincept:** Anthony Olofintuyi, Joshua Samraj, Jackelyn Samraj, Amaya Averett

**Clinical Trials Center of Middle Tennessee:** Alex Slandzicki, Jessica Wallan

**Comprehensive Pain Management and Endocrinology:** Claudia Vogel, Sebastian Munoz

**David Kavtaradze MD, Inc.:** David Kavtaradze, Casandra Watson

**David Singleton MD, PA:** David Singleton, Marcus Sevier, Maria Rivon

**DHR Health Institute for Research:** Sohail Rao, Luis Cantu

**Diabetes and Endocrinology Assoc. of Stark County:** Arvind Krishna, Heidi Daugherty, Brandi Kerr, Kathy Evans

**Doctors Medical Group of Colorado Springs, P.C.:** Robert Spees, Mailyn Marta

**Duke University:** G. Michael Felker, Meaghan Beauchaine, Amanda Harrington, Daniah Amir, Juan Collazo

**Duke University Hospital:** Rowena Dolor, Lorraine Vergara, Jackie Jordan

**Elite Family Practice:** Valencia Burruss, Terri Hurst

**Emory University:** Paulina A. Rebolledo Esteinou, Igho Ofotokun, Cecilia Zhang, Jessica Traenkner, Mary M. Atha

**Essential Medical Care, Inc.:** Vickie James, Marcella Rogers

**Family Practice Doctors P.A.:** Chukwuemeka Oragwu, Ngozi Oguego

**First Care Medical Clinic:** Rajesh Pillai, Santia Lima

**Focus Clinical Research Solutions:** Ahab Gabriel, Emad Ghaly, Marian Michal

**G&S Medical Associates, LLC:** Gammal Hassanien, Samah Ismail, Yehia Samir

**George Washington University Hospital:** Andrew Meltzer, Ryan S. Heidish, Aditya Loganathan

**Geriatrics and Medical Associates:** Scott Brehaut, Angelina Roche

**GFC of Southeastern Michigan, PC:** Manisha Mehta, Nicole Koppinger

**Health Quality Primary Care:** Jose Baez, Ivone Pagan

**Highlands Medical Associates, P.A.:** Dallal Abdelsayed, Mina Aziz

**Hoag Memorial Hospital Presbyterian**: Philip Robinson, Grace Lozinski, Julie Nguyen

**HOPE Clinical Research:** Alvin Griffin, Michael Morris, Nicole Love, Bonnie Mattox, Raykel Martin

**Hugo Medical Clinic:** Victoria Pardue, Teddy Rowland

**Innovation Clinical Trials Inc.:** Juan Ruiz-Unger, Lionel Reyes, Navila Bacallao

**Jackson Memorial Hospital:** John Cienki

**Jadestone Clinical Research, LLC:** Jonathan Cohen, Ying Yuan, Jenny Li

**Jeremy W. Szeto, D.O., P.A.:** Jeremy Szeto

**Johns Hopkins University:** Mark Sulkowski, Lauren Stelmash, Sara Mekhael

**L&A Morales Healthcare, Inc:** Idania Garcia del Sol, Ledular Morales Castillo, Anya Gutierrez, Sabrina Prieto

**Lakeland Regional Medical Center:** Arch Amon, Andrew Barbera, Andrew Bugajski, Walter Wills, Kellcee Jacklin

**Lamb Health, LLC:** Deryl Lamb, Amron Harper

**Lapis Clinical Research:** Elmer Stout, Merischia Griffin

**Loyola University Medical Center:** Nina Clark, Mary Barsanti-Sekhar, Christina Carbrera-Mendez, Mary Rose Evans

**Maria Medical Center, PLLC:** Josette Maria, Oksana Raymond

**Medical Specialists of Knoxville:** Jeffrey Summers, Tammy Turner

**Medical University of South Carolina:** Leslie Lenert, Ebony Panaccione

**Miller Family Practice, LLC:** Conrad Miller, Hawa Wiley

**Morehouse School of Medicine:** Austin Chan, Saadia Khizer

**North Shore University Health System/Evanston Hospital:** Nirav Shah, Oluwadamilola Adeyemi, Wei Ning Chi, July Chen, Melissa Morton-Jost

**Ochsner Clinic Foundation:** Julie Castex, Ali Quirch

**Olive View – UCLA Medical Center:** Hrishikesh Belani, Rosario Machicado, Bjorn Bjornsson

**Olivo Wellness Medical Center:** Jacqueline Olivo, Maria Maldonado

**Pine Ridge Family Medicine Inc.:** Anthony Vecchiarelli, Diana Gaytan-Alvarez

**Premier Health:** Vijaya Cherukuri, Santia Lima

**Providence Regional Medical Center:** Radica Alicic, Allison A. Lambert, Carissa Urbat, Joni Baxter, Ann Cooper

**Rapha Family Wellness:** Dawn Linn, Laura Fisher

**Raritan Bay Primary Care & Cardiology Associates:** Vijay Patel, Yuti Patel, Roshan Talati, Priti Patel

**Romancare Health Services:** Leonard Ellison, Angee Roman, Jeffrey Harrison

**Rush University Medical Center:** James Moy, Dina Naquiallah

**Spinal Pain and Medical Rehab, PC:** Binod Shah, Santia Lima

**Stanford University:** Prasanna Jagannathan, Upinder Singh, Orlando Quintero, Jake Scott, Andrew O’Donnell, Yasmin Jazayeri

**Sunshine Walk In Clinic:** Anita Gupta, N. Chandrasekar

**Superior Clinical Research:** Clifford Curtis, Briana White, Martha Dockery

**Tabitha B. Fortt, M.D., LLC:** Tabitha Fortt, Anisa Fortt

**Tampa General Hospital:** Jason Wilson, Jackie Marcelin, Brenda Farlow

**Temple University Hospital:** Nina Gentile, Casey Grady

**Texas Health Physicians Group:** Randall Richwine, Penny Pazier

**Texas Tech University Health Sciences Center in El Paso:** Edward Michelson, Susan Watts, Diluma Kariyawasam, Leann Rodriguez

**The Angel Medical Research:** Jose Luis Garcia, Ismarys Manresa, Angel A. Achong, Mari C. Garcia

**Trident Health Center:** Arvind Mahadevan, Santia Lima

**UF Health Precision Health Research:** Carla VandeWeerd, Jeffrey Lowenkron, Erica Sappington, Mitchell Roberts

**UMass Memorial Medical Center:** Jennifer Wang, Melissa Adams, Xinyi Ding, Mary Co

**University Diagnostics and Treatment Clinic:** Mark D'Andrea, Mina Aziz

**University Medical Center- New Orleans:** Stephen Lim, Madeline Young

**University of Arkansas:** Michael Wilson, Carly Eastin, Allyson Cheathem, Ahad Nadeem, Crystal Walters

**University of Cincinnati:** Margaret Powers-Fletcher, Douglas Brown, Delia Miller, Sylvere Mukunzi

**University of Florida-JAX-ASCENT:** Carmen Isache, Jennifer Bowman, Debra Martin

**University of Kansas – Wichita:** Brent Duran, Tiffany Schwasinger-Schmidt, Ashley Ast, Ashlie Cornejo, Allie Archer

**University of Miami:** Dushyantha Jayaweera, Maria Almanzar, Vanessa Motel

**University of Minnesota:** Matt Pullen, Neeta Bhat, Daniela Parra, Erica Urbina, Steven Arriaza

**University of Pittsburgh:** Akira Sekikawa, Emily Klawson, Jonathan Arnold, Nathan Weiland

**University of Texas Health Science Center at Houston:** Luis Ostrosky-Zeichner, Bela Patel, Virginia Umana, Laura Nielsen, Carolyn Z. Grimes

**University of Texas Health Science Center at San Antonio:** Thomas F. Patterson, Bridgette T. Soileau

**University of Virginia Health System:** Patrick E. H. Jackson, Heather M. Haughey

**Vaidya MD PLLC:** Bhavna Vaidya-Tank, Cameron Gould

**Vanderbilt University Medical Center:** Parul Goyal, Haley Pangburn, Lori Michalowski

**Wake Forest University Health Sciences:** John Williamson, Brittany Wortham, Rica Abbott

**Weill Cornell Medical College:** Matthew McCarthy, Unwana Umana, Candace Alleyne, Britta Witting

**Well Pharma Medical Research:** Eddie Armas, Ramon O. Perez Landaburo, Michelle De La Cruz, Martha Ballmajo, Jorge Alvarez

**Supplemental Methods**

**Participant Monitoring**

The daily and follow-up assessments were monitored, and sites were actively notified of events requiring review, including serious adverse events (SAEs). In addition, participants were invited during assessments to request contact from the study team or to report any unusual circumstances. Failure to complete daily assessments also triggered a review for any possible SAEs. A missed assessment on the day after receiving the first dose of study medication (day 2) or any day of missed assessments up to day 14 prompted a notification to the site to contact the participant. All participants were instructed to self-report concerns either via an online event reporting system, by calling the site, or by calling a 24-hour hotline. Hospitalizations, a record of seeking other healthcare, or serious adverse events were extracted by site personnel from the participant’s medical record. Medical occurrences occurring before the receipt of study drug/placebo but after obtaining informed consent were not considered an adverse event.

#### **Independent Data Monitoring Committee Oversight**

The chair and/or convened committee reviewed data at least once monthly to review safety data, starting when the first participants met the 28-day timepoint:

| 10/19/2023 | full DSMB review |
| --- | --- |
| 10/31/2023 | Chair Review |
| 11/28/2023 | Chair Review |
| 12/21/2023 | Chair |
| 1/9/2024 | Chair Review |
| 1/29/2024 | Full |
| 2/20/2024 | Chair review |
| 3/5/2024 | Chair |
| 3/26/2024 | Full DSMB |
| 4/2/2024 | Chair |
| 4/24/2024 | Full DSMB |
| 5/14/2024 | Chair. |
| 6/11/2024 | Chair. |
| 7/9/2024 | Chair. |
| 7/16/2024 | Full DSMB: |
| 8/06/2024 | Chair review |
| 8/27/2024 | Chair |
| 9/19/2024 | Chair |
| 10/01/2024 | Chair review |
| 11/12/2024 | Chair review |
| 12/10/2024 | Chair review |

### **Overview of Agents and Enrollment Dates on the ACTIV-6 Platform**

1. Ivermectin 400 μg/kg daily for 3 days (June 23, 2021–February 4, 2022).^1^
2. Fluvoxamine 50 mg twice daily for 10 days (August 6, 2021–May 27, 2022).^2^
3. Inhaled fluticasone furoate 200 μg daily for 14 days (August 6, 2021–February 9, 2022).^3^
4. Ivermectin 600 μg/kg daily for 6 days (February 16, 2022–July 22, 2022).^4^
5. Fluvoxamine 50 mg twice daily on day 1 followed by 100 mg twice daily for 12 days (August 25, 2022–January 20, 2023).^5^
6. Montelukast 10 mg once daily for 14 days (January 27, 2023–June 23, 2023).^6^
7. Metformin (September 19, 2023–May 1, 2024).*

- 500 mg once per day for 1 day.
- Then 500 mg in the AM and 500 mg in the PM for 4 days.
- Then 500 mg in in the AM and 1000 mg in the PM for 9 days, 14 days total (36 tablets).
- Recommend taking at the end of a small snack or meal.

*The SARS-CoV-2 JN-1 variant dominated during most of the metformin enrollment period.^7^ The L455S mutation in the receptor binding domain, the characteristic feature of the SARS-CoV-2 JN-1 variant, enhanced its infectivity, stability, and immune evasion compared to prior variants.^8,9^ Overall, however, the JN-1 variants were associated with lower severity of symptoms and reduced risk of hospitalization.^9^ Compared to the JN-1 variants, the subsequent FLiRT variants, characterized by their R346T and F456L binding domain mutations, had improved immune evasion but reduced infectivity.^10,11^ Interestingly, the R346T mutation, which increases the affinity of the viral spike protein for binding to the human ACE2, thereby enhancing immune evasion, is similar to that found in earlier Omicron subvariants.^12,13^

### **eTable 1. PASCD treatment effect estimates with limited covariate adjustment**

| **Day** | **Metformin** | **Placebo** | **Delta** | **PPE** | **Ratio** |
| --- | --- | --- | --- | --- | --- |
| 90 | 0.038 (0.028, 0.050) | 0.041 (0.031, 0.053) | -0.003 (-0.019, 0.013) | 0.655 | 0.921 (0.588, 1.337) |
| 120 | 0.035 (0.025, 0.046) | 0.047 (0.036, 0.058) | -0.012 (-0.027, 0.003) | 0.935 | 0.739 (0.495, 1.063) |
| 180 | 0.028 (0.019, 0.038) | 0.036 (0.026, 0.046) | -0.008 (-0.022, 0.006) | 0.849 | 0.784 (0.476, 1.184) |
| 90, 120, or 180 | 0.073 (0.060, 0.087) | 0.086 (0.072, 0.101) | -0.013 (-0.032, 0.007) | 0.891 | 0.852 (0.663, 1.083) |

Delta = Risk difference

PPE = Posterior probability of efficacy

Ratio = Risk ratio

### Adjusted for BMI (indicator for greater than 30), prior COVID-19 infection, and vaccination status

### **eTable 2. Baseline characteristics between those who responded to at least 1 long-term follow-up survey versus those who did not**

| **Variable** | **At least one long-term survey response** | **No long-term survey response** | **Overall** |
| --- | --- | --- | --- |
|  | 2563 | 420 | 2983 |
| Age, median (IQR), y | 48.0 (38.0-58.0) | 45.0 (36.0-57.0) | 47.0 (38.0-58.0) |
| Age < 50, No. (%) | 1387 (54.1) | 252 (60.0) | 1639 (54.9) |
| Biologic sex at birth, No. (%) |  |  |  |
| Female | 1619 (63.17) | 271 (64.52) | 1890 (63.36) |
| Male | 937 (36.56) | 148 (35.24) | 1085 (36.37) |
| Undifferentiated | 2 (0.08) | 0 (0.00) | 2 (0.07) |
| Unknown | 2 (0.08) | 1 (0.24) | 3 (0.10) |
| Prefer not to answer | 3 (0.12) | 0 (0.00) | 3 (0.10) |
| Race^a^, No (%) |  |  |  |
| American Indian or Alaska Native | 20 (0.78) | 5 (1.19) | 25 (0.84) |
| Asian | 64 (2.50) | 13 (3.10) | 77 (2.58) |
| Black or African American | 278 (10.85) | 70 (16.67) | 348 (11.67) |
| Middle Eastern or North African | 41 (1.60) | 17 (4.05) | 58 (1.94) |
| Native Hawaiian, Other Pacific Islander | 7 (0.27) | 1 (0.24) | 8 (0.27) |
| White | 2108 (82.25) | 282 (67.14) | 2390 (80.12) |
| None of the above | 46 (1.79) | 22 (5.24) | 68 (2.28) |
| Prefer not to answer | 36 (1.40) | 17 (4.05) | 53 (1.78) |
| Ethnicity, No. (%) |  |  |  |
| Hispanic/Latino | 1253 (48.89) | 138 (32.86) | 1391 (46.63) |
| Not Hispanic/Latino | 1310 (51.11) | 282 (67.14) | 1592 (53.37) |
| Region, No. (%) |  |  |  |
| Midwest | 496 (19.35) | 107 (25.48) | 603 (20.21) |
| Northeast | 190 (7.41) | 60 (14.29) | 250 (8.38) |
| South | 1621 (63.25) | 209 (49.76) | 1830 (61.35) |
| West | 256 (9.99) | 44 (10.48) | 300 (10.06) |
| Call center, No. (%) | 78 (3.04) | 5 (1.19) | 83 (2.78) |
| BMI, median (IQR), kg/m^2 | 28.3 (25.2-32.1) | 29.0 (25.6-32.9) | 28.4 (25.2-32.2) |
| BMI > 30 kg/m^2, No. (%) | 963 (37.6) | 176 (41.9) | 1139 (38.2) |
| Weight > 88 kg, No. (%) | 825 (32.2) | 153 (36.4) | 978 (32.8) |
| Heart disease, No./total (%) | 75/2520 (2.98) | 14/341 (4.11) | 89/2861 (3.11) |
| Diabetes, No./total (%) | 42/2520 (1.67) | 15/341 (4.40) | 57/2861 (1.99) |
| High blood pressure, No./total (%) | 521/2520 (20.67) | 76/341 (22.29) | 597/2861 (20.87) |
| COPD, No./total (%) | 51/2520 (2.02) | 7/341 (2.05) | 58/2861 (2.03) |
| Asthma, No./total (%) | 234/2520 (9.29) | 42/341 (12.32) | 276/2861 (9.65) |
| Chronic kidney disease, No./total (%) | 7/2520 (0.28) | 0/341 (0.00) | 7/2861 (0.24) |
| Smoker, past year, No./total (%) | 371/2520 (14.72) | 54/341 (15.84) | 425/2861 (14.85) |
| Malignant cancer, No./total (%) | 57/2515 (2.27) | 3/339 (0.88) | 60/2854 (2.10) |
| Prior COVID-19 infectino, No. (%) | 1414 (55.17) | 203 (48.33) | 1617 (54.21) |
| Vaccine status, No. (%) |  |  |  |
| Not vaccinated | 759 (29.61) | 182 (43.33) | 941 (31.55) |
| Vaccinated (1 dose) | 4 (0.16) | 0 (0.00) | 4 (0.13) |
| Vaccinated (2+ doses) | 1800 (70.23) | 238 (56.67) | 2038 (68.32) |
| Days between symptom onset and receipt of drug, median (IQR) | 5 (4-6) | 5 (4-7) | 5 (4-7) |
| Days between symptom onset and enrollment, median (IQR) | 3 (2-5) [n=2546] | 3 (2-5) [n=370] | 3 (2-5) [n=2916] |
| Symptom burden on/before day of drug receipt, No. (%) | | | |
| None | 102 (4.0) | 36 (8.6) | 138 (4.6) |
| Mild | 1145 (44.7) | 191 (45.5) | 1336 (44.8) |
| Moderate | 1244 (48.5) | 168 (40.0) | 1412 (47.3) |
| Severe | 72 (2.8) | 25 (6.0) | 97 (3.3) |
| Remdesivir, No. (%) | 0 (0.00) | 1 (0.24) | 1 (0.03) |
| Nirmatrelvir/ritonavir, No. (%) | 371 (14.48) | 62 (14.76) | 433 (14.52) |
| Monoclonal antibodies, No. (%) | 61 (2.38) | 13 (3.10) | 74 (2.48) |
| Molnupiravir, No. (%) | 21 (0.82) | 2 (0.48) | 23 (0.77) |

Participants may have selected any combination of the following race descriptors: American Indian or Alaska Native; Asian; Black, African American, or African; Middle Eastern or North African; Native Hawaiian or Other Pacific Islander; White; None of the above; Prefer not to answer

### **eTable 3. Outcome counts across all time points**

| **Outcome** | **Day** | **Metformin**  **n=1439** | | | | **Placebo**  **n=1544** | | | |
| --- | --- | --- | --- | --- | --- | --- | --- | --- | --- |
|  |  | **Events** | **n*** | **(%)** | **% missing** | **Events** | **n*** | **(%)** | **% missing** |
| PASCD | 90, 120, 180 | 117 | 1234 | 9.48 | 14.3 | 159 | 1329 | 11.96 | 13.9 |
|  |  | 117 | 1439 | 8.13 |  | 159 | 1544 | 10.30 |  |
|  | 90, 120, 180 | 71 | 1234 | 5.75 | 14.3 | 97 | 1329 | 7.30 | 13.9 |
|  |  | 71 | 1439 | 4.93 |  | 97 | 1544 | 6.28 |  |
| New Infection | 120, 180 | 12 | 1234 | 0.97 | 14.3 | 11 | 1329 | 0.83 | 13.9 |
|  |  | 12 | 1439 | 0.83 |  | 11 | 1544 | 0.71 |  |

*The top row is among those who filled out at least one survey on or after day 90 and the bottom row is among the mITT.

### **eTable 4. Symptom burden and new infection responses at each time point**

|  | **Metformin (N = 1439)** | **Placebo (N = 1544)** | **Overall (N=2983)** |
| --- | --- | --- | --- |
| **Symptom burden**, No. (%) |  |  |  |
| Day 90 |  |  |  |
| None | 1119 (77.8) | 1216 (78.8) | 2335 (78.3) |
| Mild | 39 (2.7) | 40 (2.6) | 79 (2.6) |
| Moderate | 5 (0.3) | 12 (0.8) | 17 (0.6) |
| Severe | 0 (0.0) | 0 (0.0) | 0 (0.0) |
| Missing | 276 (19.2) | 276 (17.9) | 552 (18.5) |
| Day 120 |  |  |  |
| None | 1118 (77.7) | 1193 (77.3) | 2311 (77.5) |
| Mild | 33 (2.3) | 56 (3.6) | 89 (3.0) |
| Moderate | 8 (0.6) | 9 (0.6) | 17 (0.6) |
| Severe | 0 (0.0) | 0 (0.0) | 0 (0.0) |
| Missing | 280 (19.5) | 286 (18.5) | 566 (19.0) |
| Day 180 |  |  |  |
| None | 1101 (76.5) | 1203 (77.9) | 2304 (77.2) |
| Mild | 27 (1.9) | 41 (2.7) | 68 (2.3) |
| Moderate | 11 (0.8) | 7 (0.5) | 18 (0.6) |
| Severe | 0 (0.0) | 0 (0.0) | 0 (0.0) |
| Missing | 300 (20.8) | 293 (19.0) | 593 (19.9) |
| **New COVID infection within 28 days prior to the follow-up survey**, No. (%) | | | |
| Day 120 |  |  |  |
| Yes | 2 (0.1) | 6 (0.4) | 8 (0.3) |
| No | 1157 (80.4) | 1252 (81.1) | 2409 (80.8) |
| Missing | 280 (19.5) | 286 (18.5) | 566 (19.0) |
| Day 180 |  |  |  |
| Yes | 10 (0.7) | 5 (0.3) | 15 (0.5) |
| No | 1129 (78.5) | 1246 (80.7) | 2375 (79.6) |
| Missing | 300 (20.8) | 293 (19.0) | 593 (19.9) |

### **eTable 5. Observed counts by survey day and baseline characteristic, clinician diagnosis of long COVID**

| **Measure** | **Variable** | **Metformin** | **Placebo** |
| --- | --- | --- | --- |
| **Day 120** |  |  |  |
| Days from symptom onset | (0,1] | 0 (0.0) | 1/7 (14.3) |
|  | (1,2] | 0/61 (0.0) | 1/68 (1.5) |
|  | (2,3] | 3/174 (1.7) | 1/198 (0.5) |
|  | (3,4] | 2/240 (0.8) | 2/236 (0.8) |
|  | (4,5] | 2/223 (0.9) | 3/257 (1.2) |
|  | (5,14] | 6/451 (1.3) | 8/491 (1.6) |
| Prior infection | Yes | 9/623 (1.4) | 8/640 (1.2) |
|  | No | 1/489 (0.2) | 7/575 (1.2) |
| Baseline symptoms | None | 0/44 (0.0) | 0/43 (0.0) |
|  | Mild | 5/509 (1.0) | 4/562 (0.7) |
|  | Moderate | 7/576 (1.2) | 10/614 (1.6) |
|  | Severe | 1/30 (3.3) | 2/38 (5.3) |
| BMI | (0,25] | 3/260 (1.2) | 3/305 (1.0) |
|  | (25,30] | 6/443 (1.4) | 5/517 (1.0) |
|  | (30,100] | 4/456 (0.9) | 8/435 (1.8) |
| **Day 180** |  |  |  |
| Days from symptom onset | (0,1] | 0 (0.0) | 1/7 (14.3) |
|  | (1,2] | 0/58 (0.0) | 2/65 (3.1) |
|  | (2,3] | 1/175 (0.6) | 3/193 (1.6) |
|  | (3,4] | 2/236 (0.8) | 0/230 (0.0) |
|  | (4,5] | 1/217 (0.5) | 5/257 (1.9) |
|  | (5,14] | 4/441 (0.9) | 7/499 (1.4) |
| Prior infection | Yes | 5/614 (0.8) | 10/648 (1.5) |
|  | No | 3/479 (0.6) | 6/561 (1.1) |
| Baseline symptoms | None | 0/45 (0.0) | 1/49 (2.0) |
|  | Mild | 2/498 (0.4) | 5/560 (0.9) |
|  | Moderate | 6/568 (1.1) | 9/608 (1.5) |
|  | Severe | 0/27 (0.0) | 3/34 (8.8) |
| BMI | (0,25] | 2/260 (0.8) | 2/306 (0.7) |
|  | (25,30] | 3/430 (0.7) | 7/506 (1.4) |
|  | (30,100] | 3/448 (0.7) | 9/439 (2.1) |

### **eTable 6. Observed counts by survey day and baseline characteristic, PASCD**

| **Measure** | **Variable** | **Metformin** | **Placebo** |
| --- | --- | --- | --- |
| **Day 90** | | | |
| Days from symptom onset | (0,1] | 2 (20.0) | 2/8 (25.0) |
|  | (1,2] | 2/64 (3.1) | 4/68 (5.9) |
|  | (2,3] | 8/180 (4.4) | 5/203 (2.5) |
|  | (3,4] | 4/247 (1.6) | 8/238 (3.4) |
|  | (4,5] | 5/218 (2.3) | 11/258 (4.3) |
|  | (5,14] | 23/444 (5.2) | 22/493 (4.5) |
| Prior infection | Yes | 27/626 (4.3) | 30/650 (4.6) |
|  | No | 17/492 (3.5) | 21/572 (3.7) |
| Baseline symptoms | None | 1/42 (2.4) | 1/44 (2.3) |
|  | Mild | 15/510 (2.9) | 19/567 (3.4) |
|  | Moderate | 24/578 (4.2) | 26/621 (4.2) |
|  | Severe | 4/33 (12.1) | 6/36 (16.7) |
| BMI | (0,25] | 9/260 (3.5) | 11/305 (3.6) |
|  | (25,30] | 12/444 (2.7) | 19/519 (3.7) |
|  | (30,100] | 23/459 (5.0) | 22/444 (5.0) |
| **Day 120** | | | |
| Days from symptom onset | (0,1] | 1 (10.0) | 1/7 (14.3) |
|  | (1,2] | 1/61 (1.6) | 4/68 (5.9) |
|  | (2,3] | 5/174 (2.9) | 7/198 (3.5) |
|  | (3,4] | 7/240 (2.9) | 7/236 (3.0) |
|  | (4,5] | 5/223 (2.2) | 9/258 (3.5) |
|  | (5,14] | 21/451 (4.7) | 33/491 (6.7) |
| Prior infection | Yes | 26/623 (4.2) | 35/641 (5.5) |
|  | No | 12/489 (2.5) | 23/575 (4.0) |
| Baseline symptoms | None | 0/44 (0.0) | 1/43 (2.3) |
|  | Mild | 15/509 (2.9) | 28/562 (5.0) |
|  | Moderate | 22/576 (3.8) | 26/615 (4.2) |
|  | Severe | 3/30 (10.0) | 6/38 (15.8) |
| BMI | (0,25] | 6/260 (2.3) | 14/305 (4.6) |
|  | (25,30] | 16/443 (3.6) | 19/517 (3.7) |
|  | (30,100] | 18/456 (3.9) | 28/436 (6.4) |
| **Day 180** | | | |
| Days from symptom onset | (0,1] | 2 (20.0) | 1/7 (14.3) |
|  | (1,2] | 0/58 (0.0) | 2/65 (3.1) |
|  | (2,3] | 6/175 (3.4) | 4/193 (2.1) |
|  | (3,4] | 5/236 (2.1) | 8/230 (3.5) |
|  | (4,5] | 8/217 (3.7) | 5/257 (1.9) |
|  | (5,14] | 12/442 (2.7) | 26/499 (5.2) |
| Prior infection | Yes | 26/614 (4.2) | 22/648 (3.4) |
|  | No | 4/480 (0.8) | 20/561 (3.6) |
| Baseline symptoms | None | 0/45 (0.0) | 0/49 (0.0) |
|  | Mild | 12/499 (2.4) | 21/560 (3.8) |
|  | Moderate | 17/568 (3.0) | 20/608 (3.3) |
|  | Severe | 4/27 (14.8) | 5/34 (14.7) |
| BMI | (0,25] | 9/260 (3.5) | 9/306 (2.9) |
|  | (25,30] | 11/431 (2.6) | 19/506 (3.8) |
|  | (30,100] | 13/448 (2.9) | 18/439 (4.1) |
| **Day 90, 120, or 180 among complete responders** | | | |
| Days from symptom onset | (0,1] | 3 (30.0) | 1/6 (16.7) |
|  | (1,2] | 3/56 (5.4) | 5/59 (8.5) |
|  | (2,3] | 10/166 (6.0) | 10/188 (5.3) |
|  | (3,4] | 12/225 (5.3) | 14/223 (6.3) |
|  | (4,5] | 9/203 (4.4) | 15/243 (6.2) |
|  | (5,14] | 34/405 (8.4) | 52/455 (11.4) |
| Prior infection | Yes | 47/581 (8.1) | 54/603 (9.0) |
|  | No | 19/448 (4.2) | 37/535 (6.9) |
| Baseline symptoms | None | 0/36 (0.0) | 1/38 (2.6) |
|  | Mild | 25/462 (5.4) | 43/525 (8.2) |
|  | Moderate | 39/540 (7.2) | 47/580 (8.1) |
|  | Severe | 7/27 (25.9) | 6/31 (19.4) |
| BMI | (0,25] | 13/238 (5.5) | 20/290 (6.9) |
|  | (25,30] | 23/407 (5.7) | 36/484 (7.4) |
|  | (30,100] | 35/420 (8.3) | 41/400 (10.2) |
| Day 90, 120, 180 among complete responders | | | |

### **eFigure 1. Heterogeneity of treatment effect, day 180 PASCD**

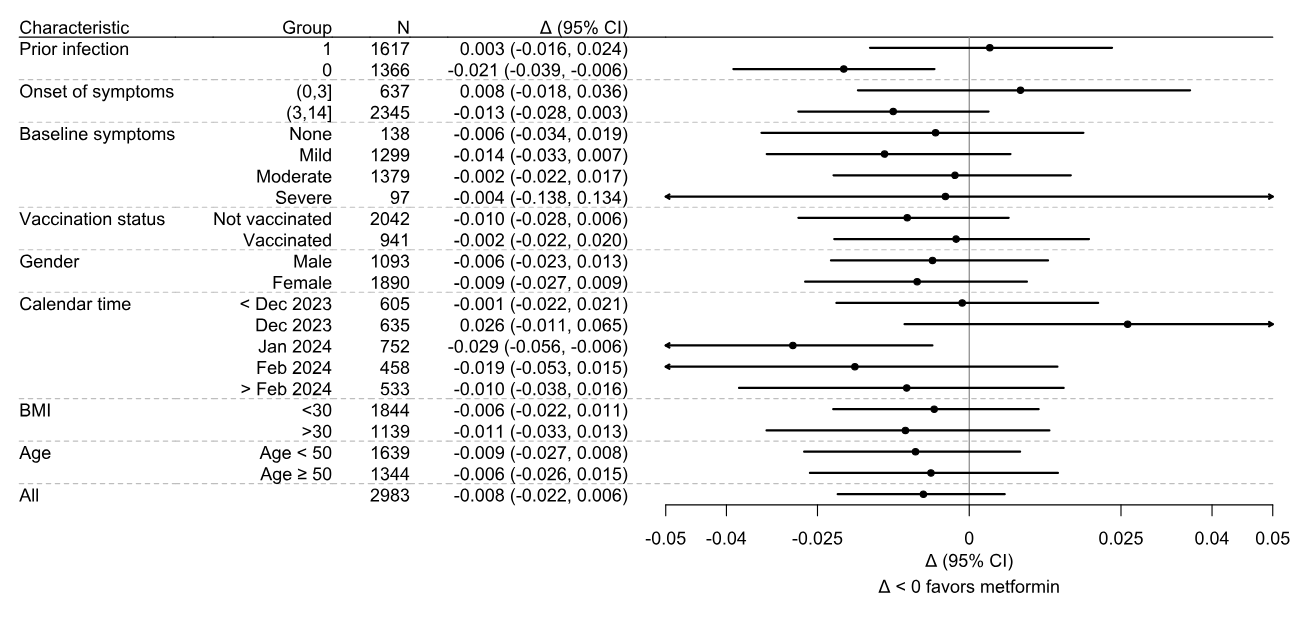

This figure reports the treatment effect estimated within different levels of a baseline characteristic. The treatment effects are calculated from a covariate adjusted model that also includes an interaction term between the treatment assignment and the characteristic of interest. The posterior distribution within each level is calculated by averaging the risk difference over all participants within the level for each draw or the model posterior. The risk difference and credible interval are calculated from the level-specific posterior distributions.

### **eFigure 2. Posterior probability distributions of PASCD on day 180, model-based estimate of treatment effect**

| 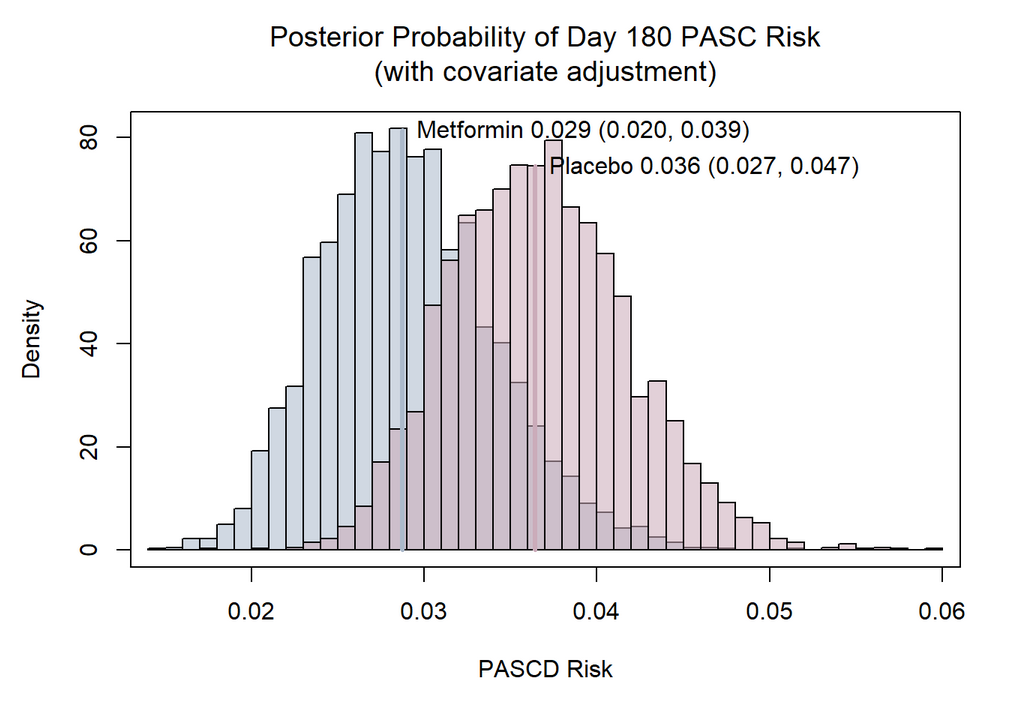 | 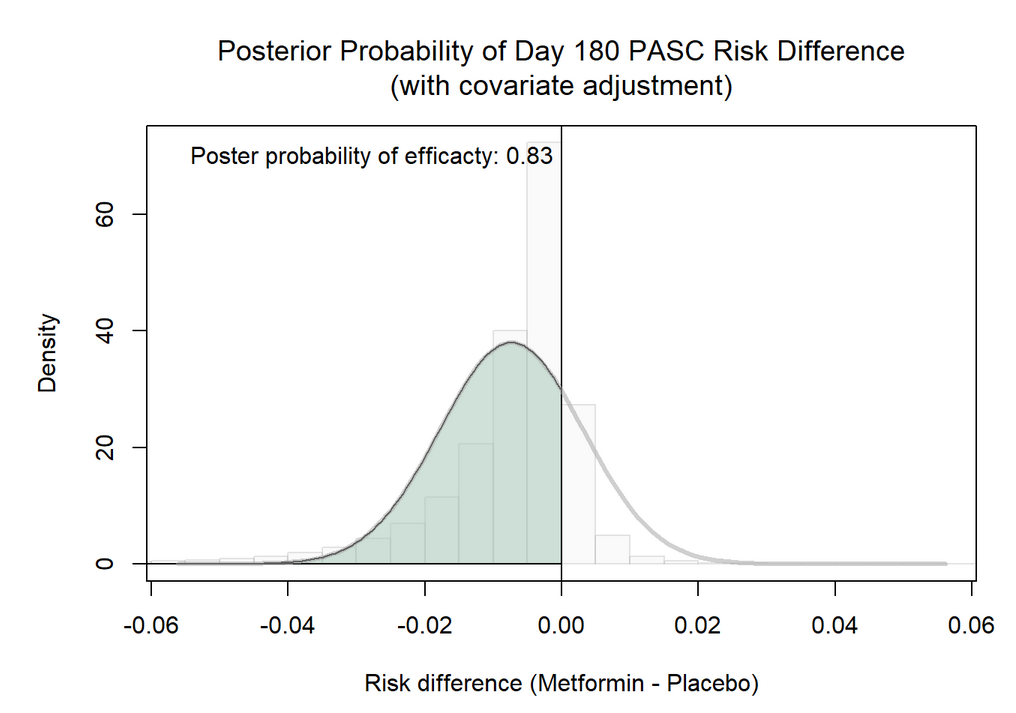 |
| --- | --- |

### **eFigure 3. Posterior probability distributions of PASCD on day 90 and 120, model-based estimate of treatment effect**

**z**

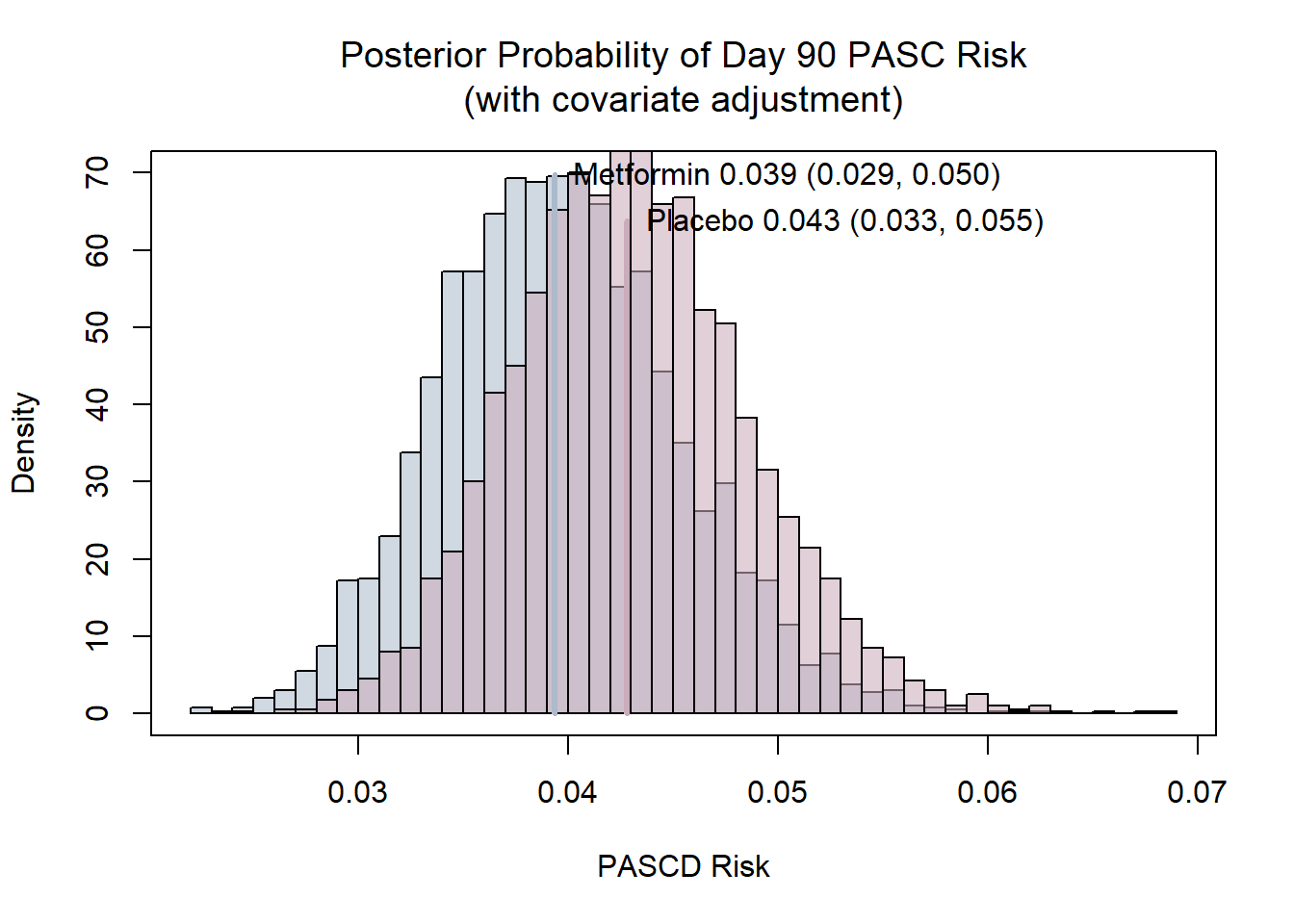

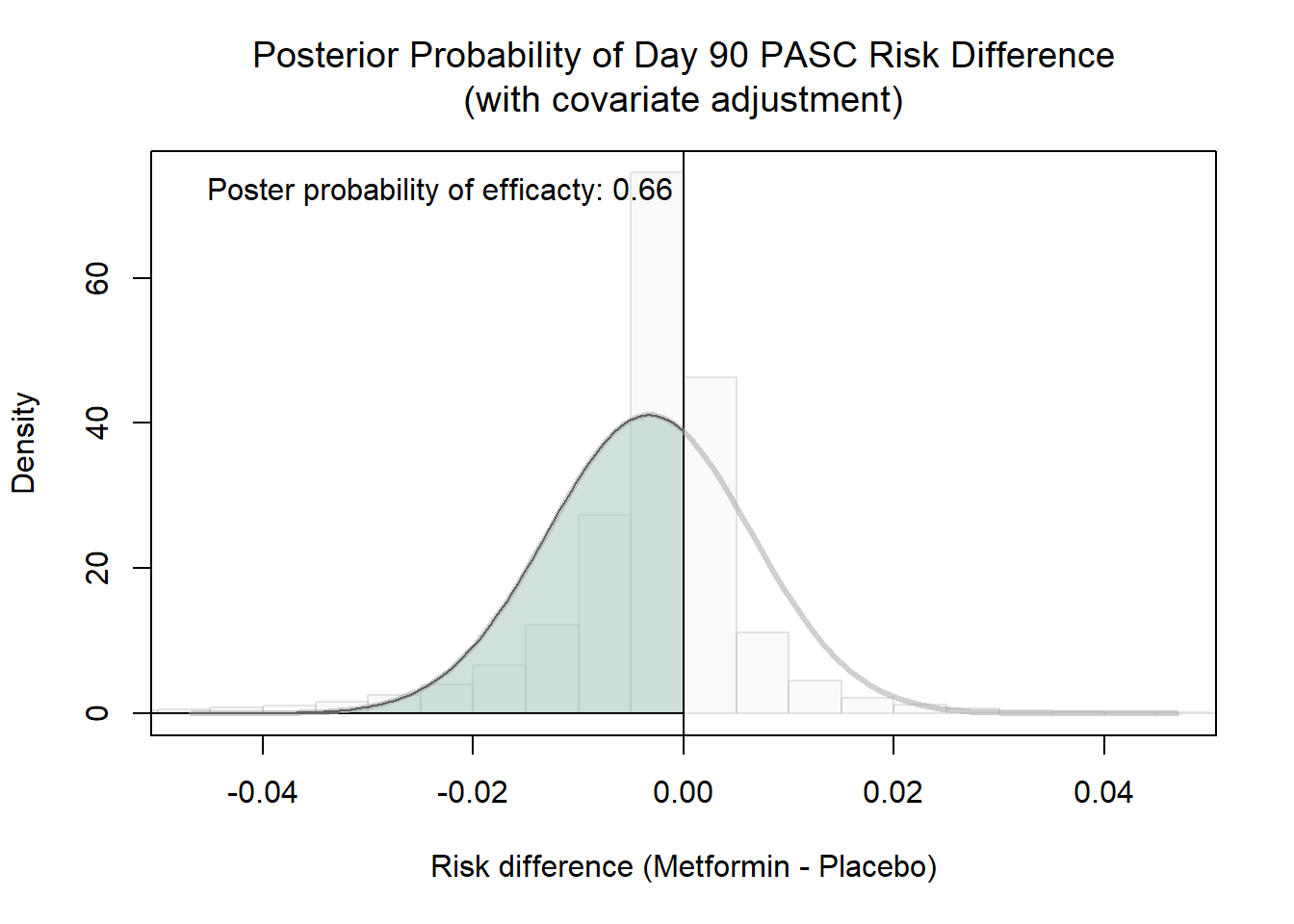

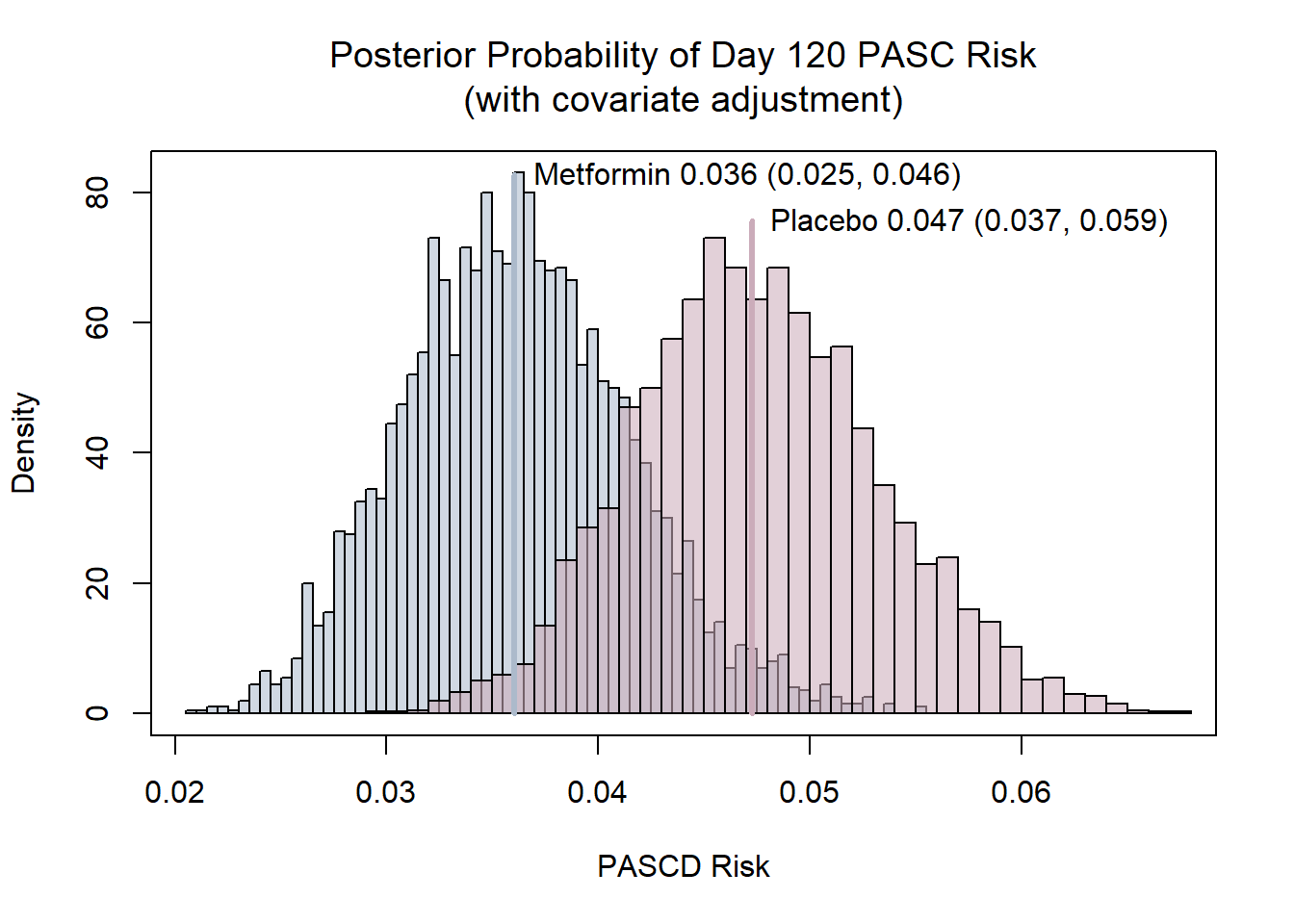

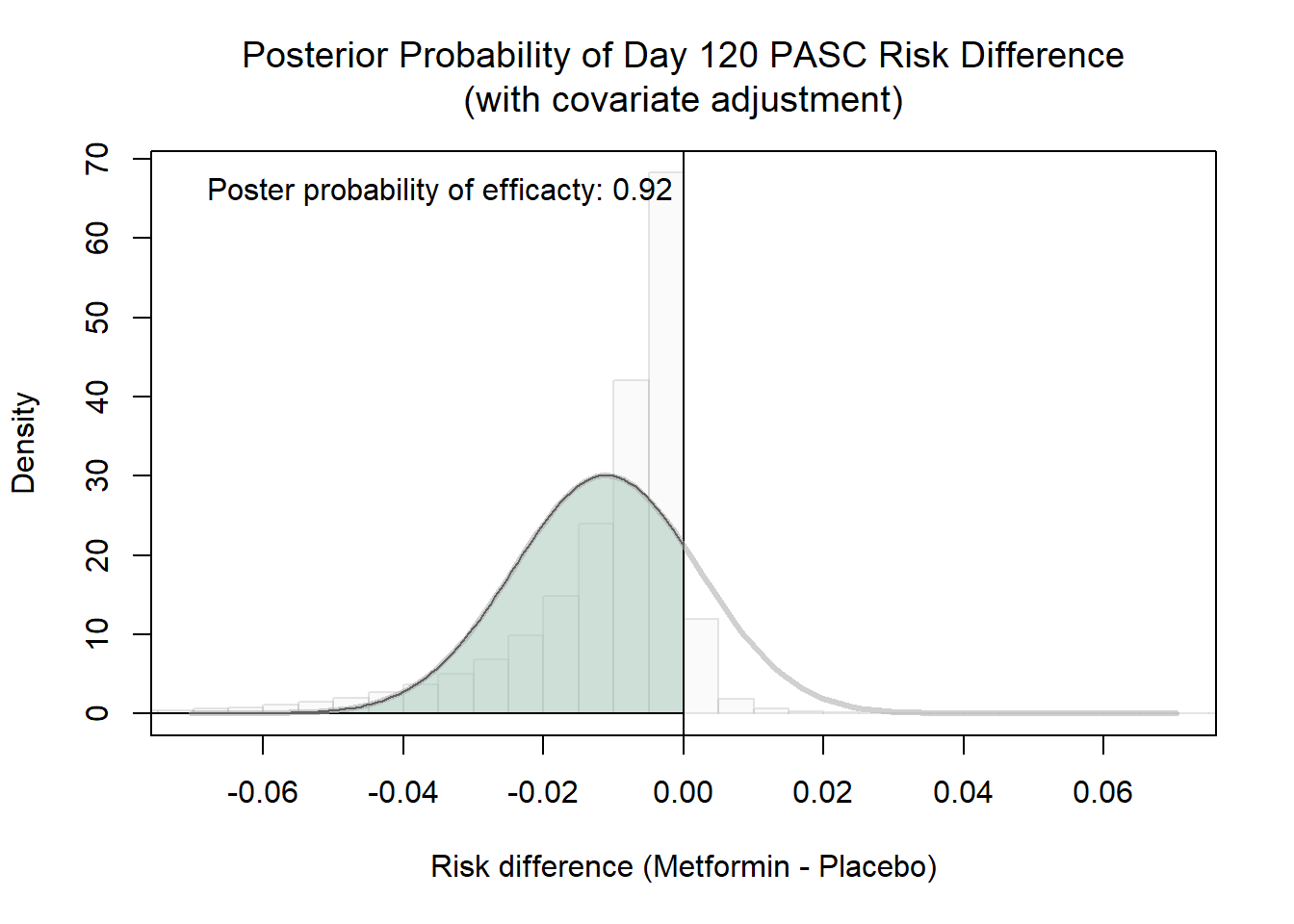

### **eFigure 4. Posterior probability distributions of PASCD relative risks, covariate adjusted.**

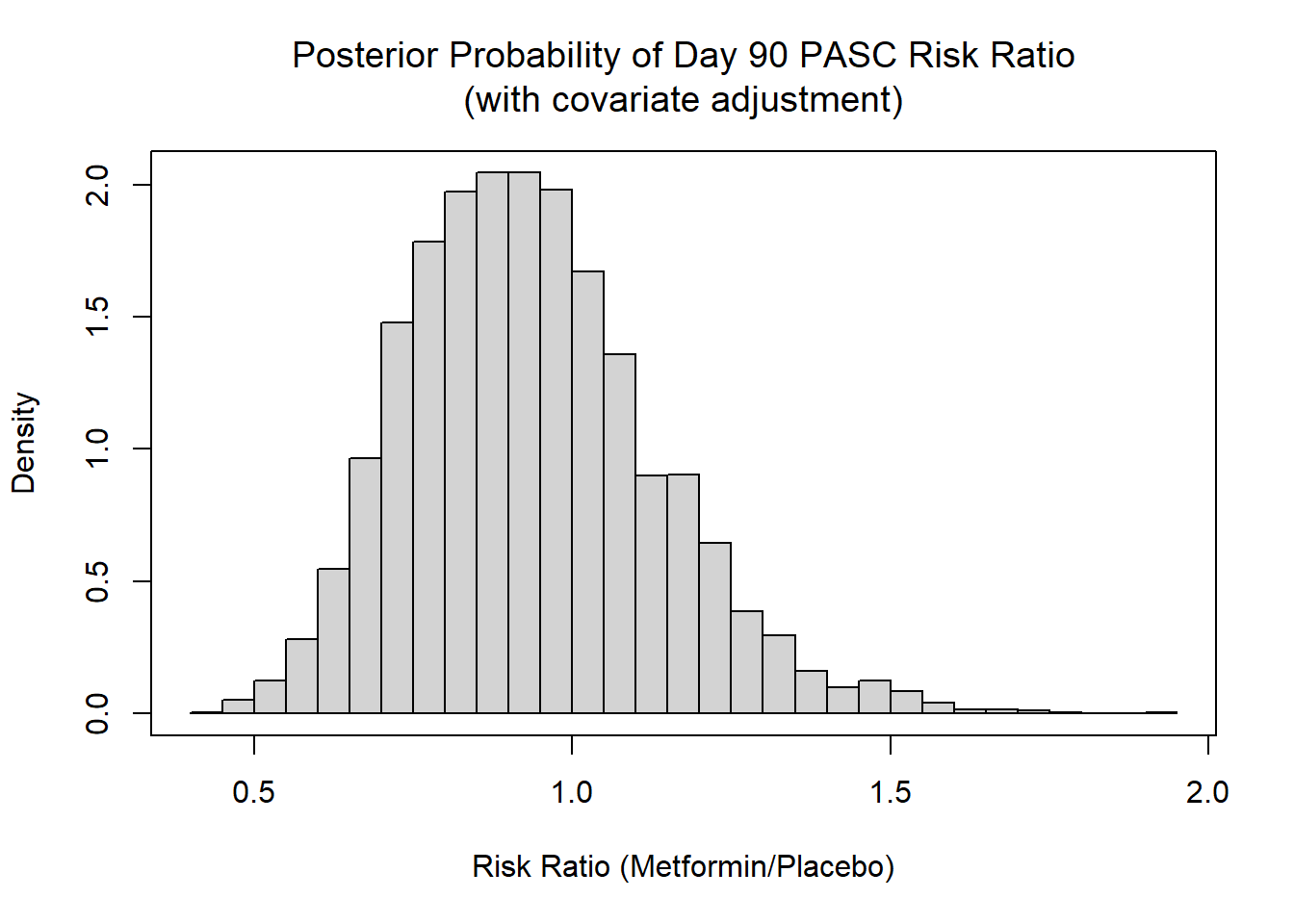

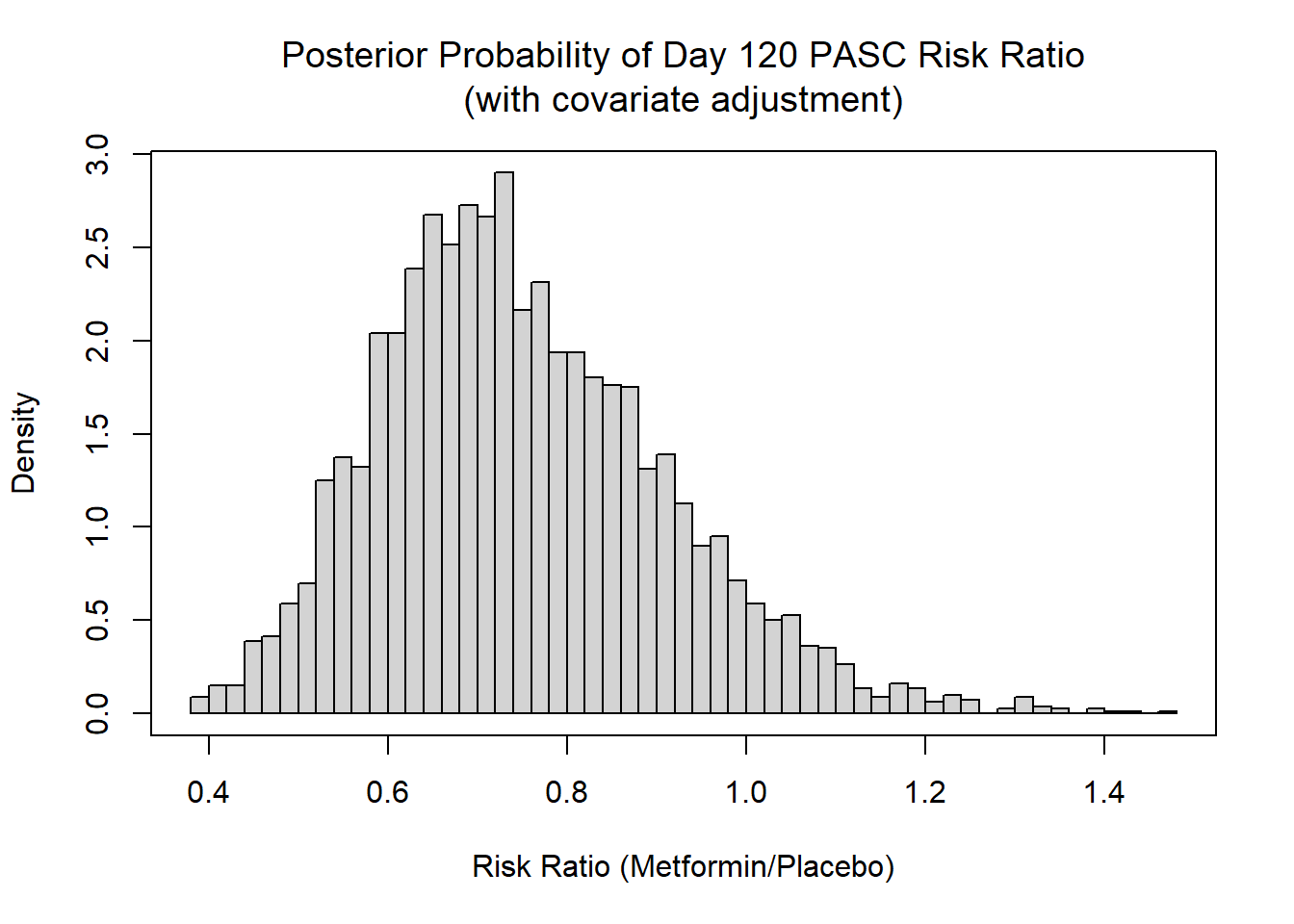

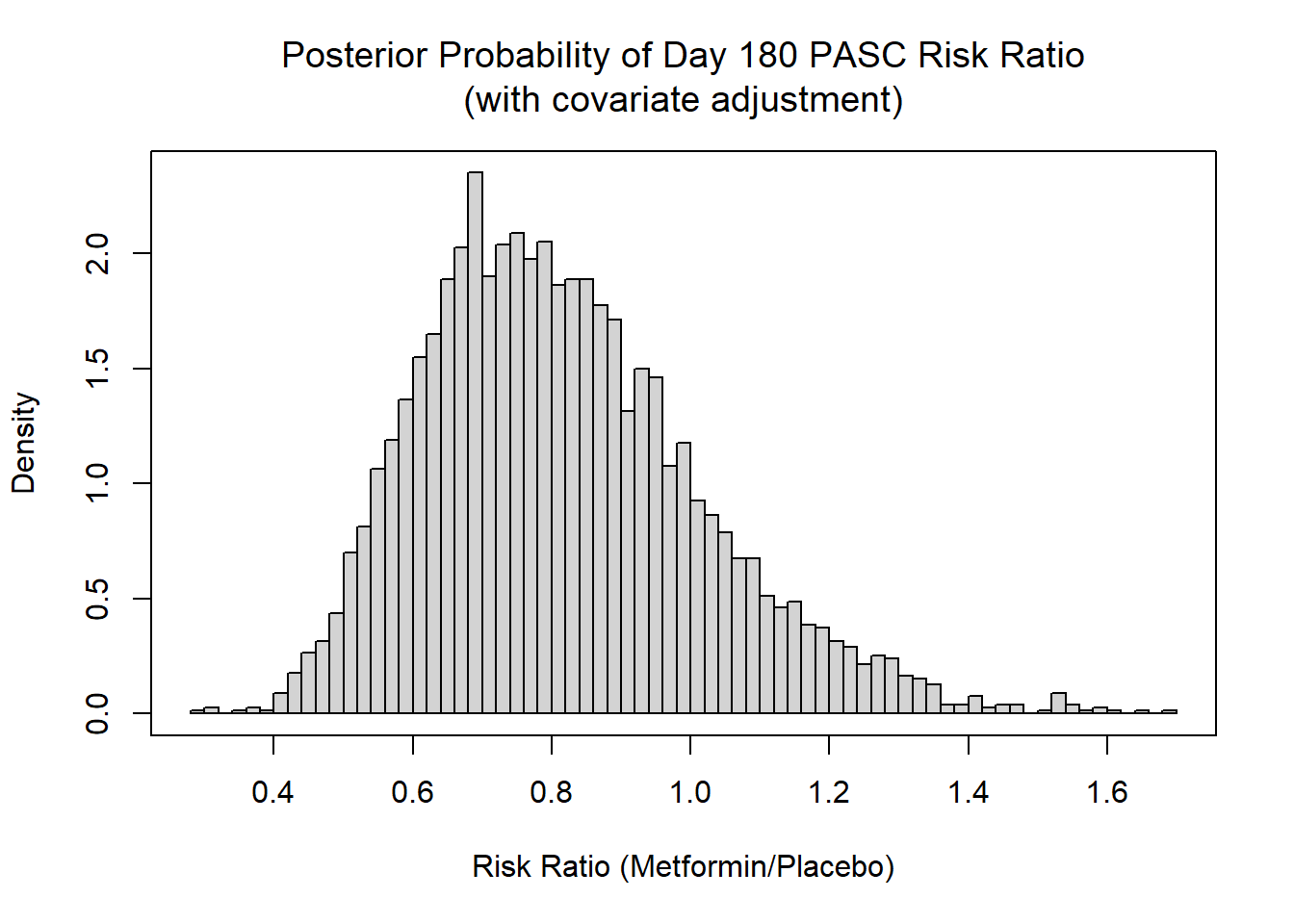

### **eFigure 5. Symptom burden, model-based estimates of treatment effect**

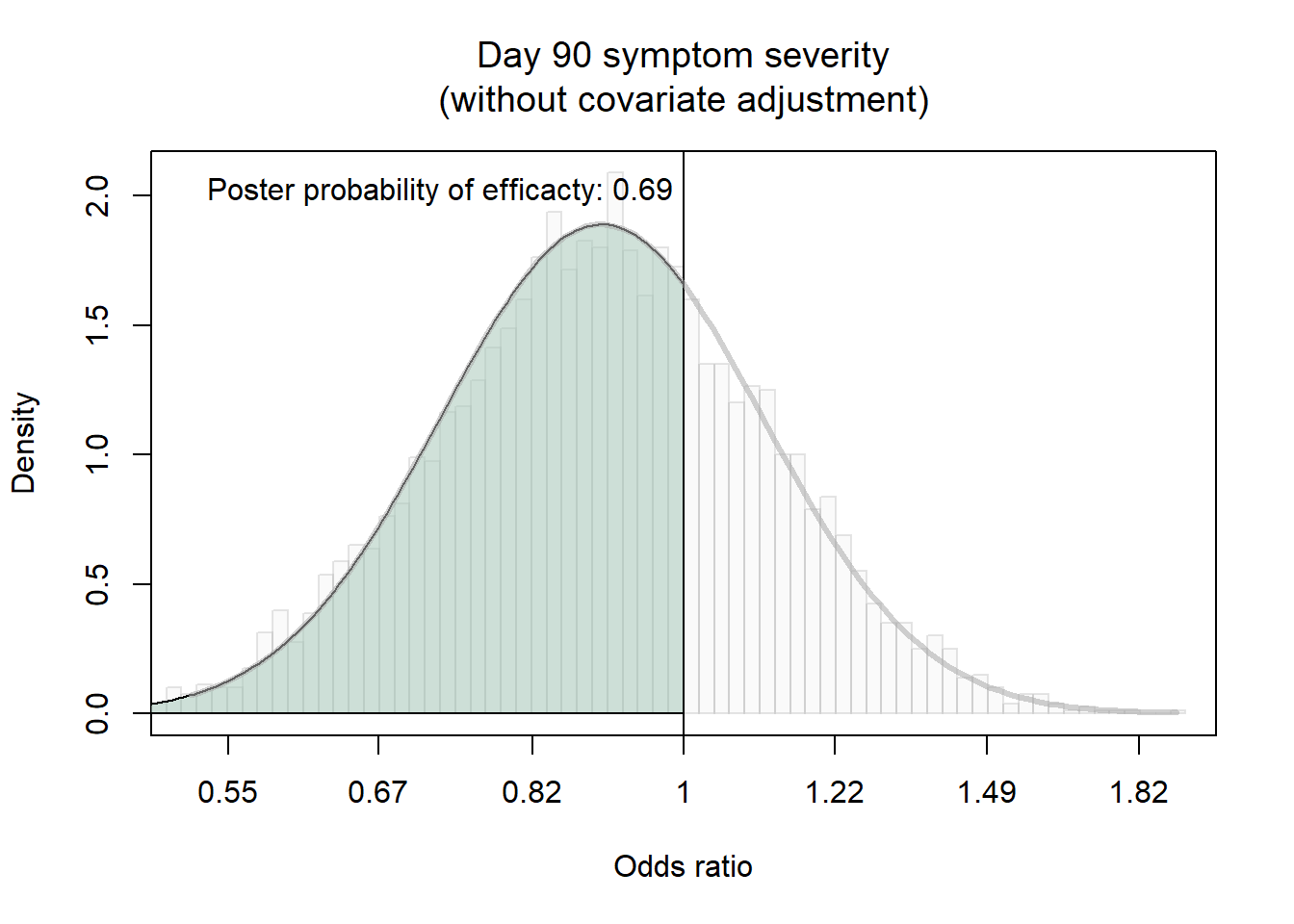

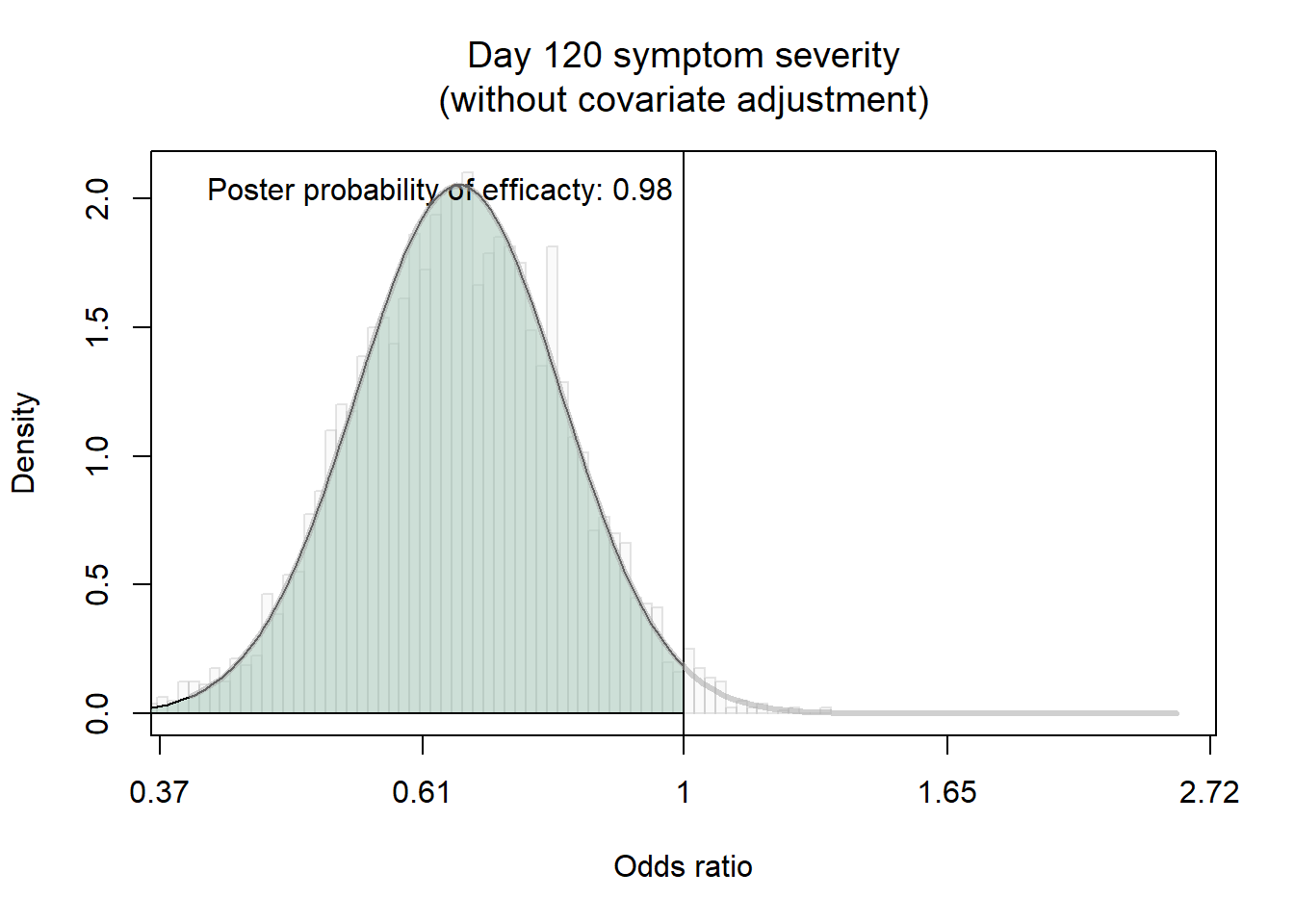

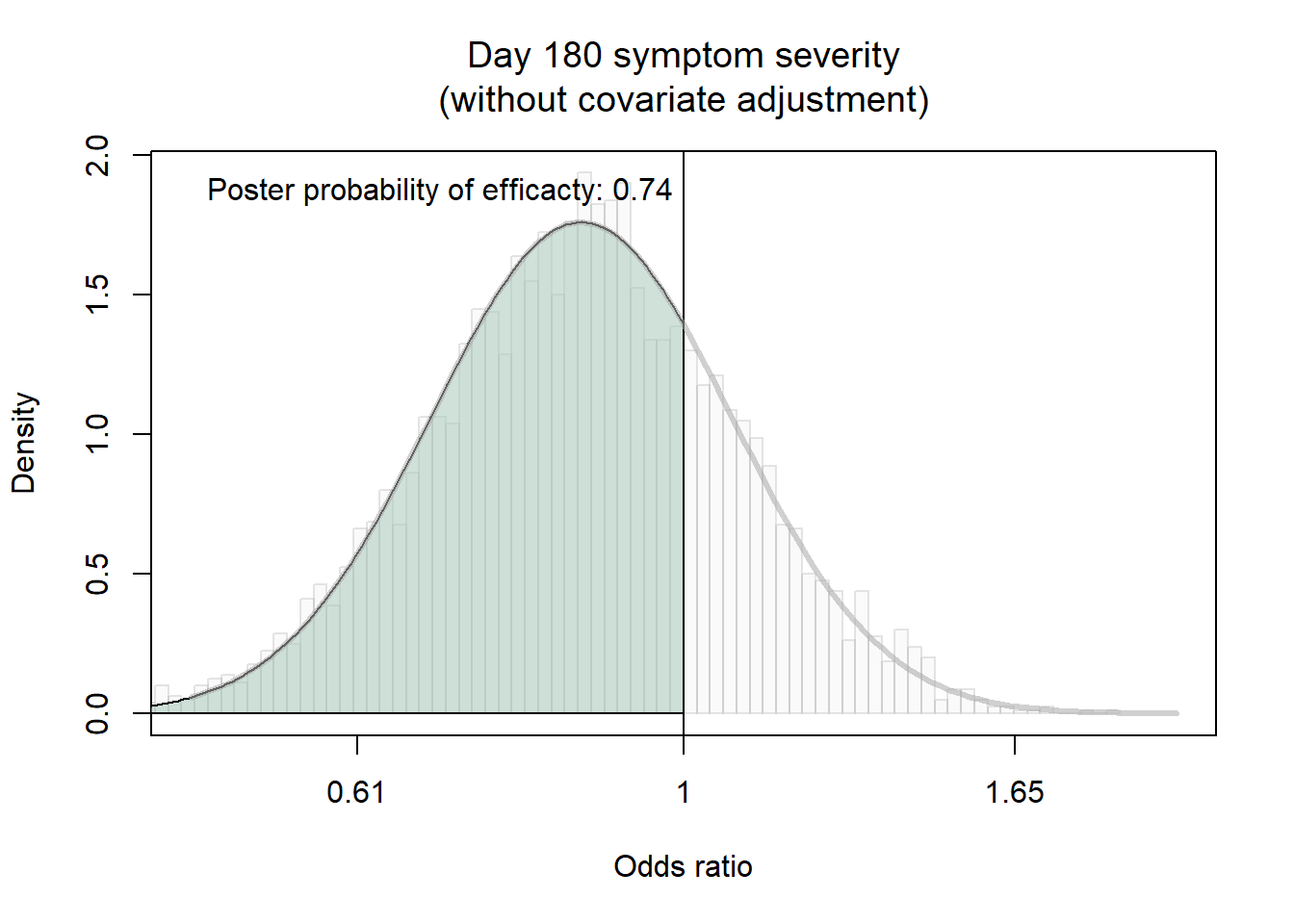

### **eFigure 6. Clinician Diagnosis of long COVID, model-based estimates of treatment effect**

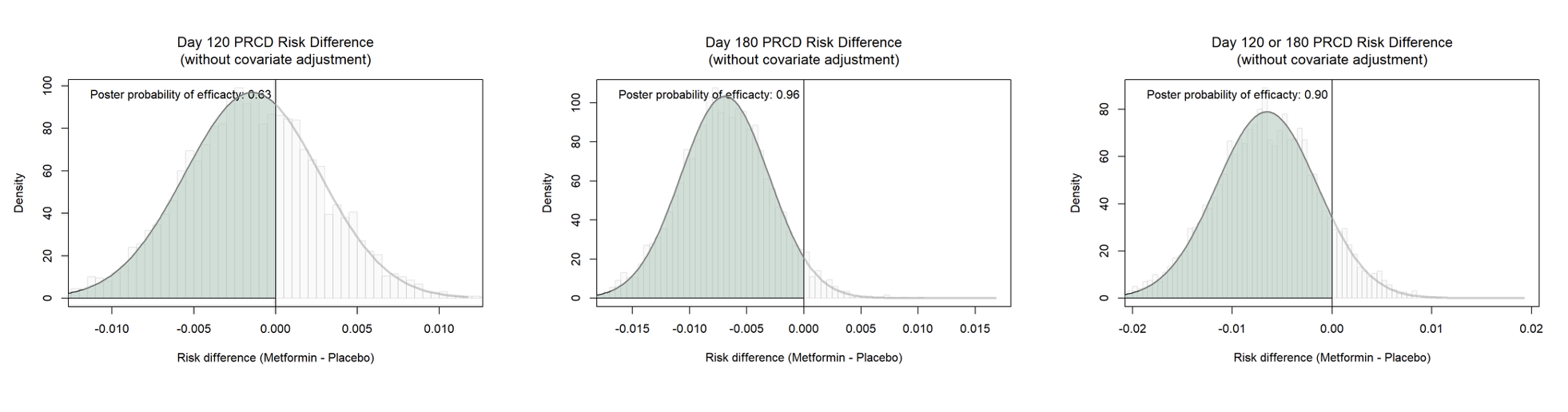

### **eFigure 7. Posterior distributions for PASCD treatment effect on day 180 comparing participants who responded to at least one long-term follow-up survey versus those who did not**

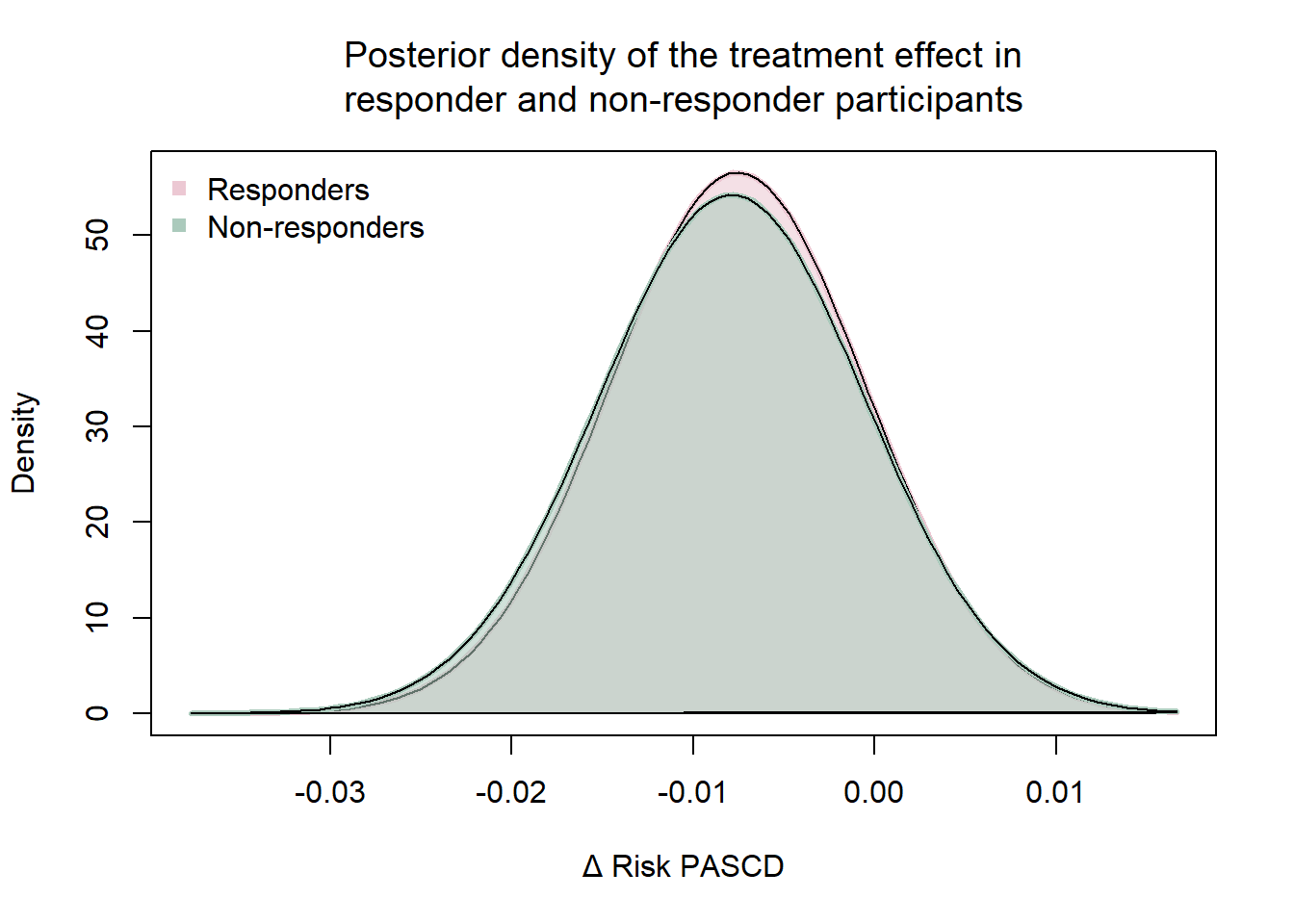

### **References**

1.	Naggie S, Boulware DR, Lindsell CJ, et al. Effect of Ivermectin vs Placebo on Time to Sustained Recovery in Outpatients With Mild to Moderate COVID-19: A Randomized Clinical Trial. *JAMA*. Oct 25 2022;328(16):1595-1603. doi:10.1001/jama.2022.18590

2.	McCarthy MW, Naggie S, Boulware DR, et al. Effect of Fluvoxamine vs Placebo on Time to Sustained Recovery in Outpatients With Mild to Moderate COVID-19: A Randomized Clinical Trial. *JAMA*. Jan 24 2023;329(4):296-305. doi:10.1001/jama.2022.24100

3.	Boulware DR, Lindsell CJ, Stewart TG, et al. Inhaled Fluticasone Furoate for Outpatient Treatment of Covid-19. *N Engl J Med*. Sep 21 2023;389(12):1085-1095. doi:10.1056/NEJMoa2209421

4.	Naggie S, Boulware DR, Lindsell CJ, et al. Effect of Higher-Dose Ivermectin for 6 Days vs Placebo on Time to Sustained Recovery in Outpatients With COVID-19: A Randomized Clinical Trial. *JAMA*. Mar 21 2023;329(11):888-897. doi:10.1001/jama.2023.1650

5.	Accelerating C-TI, Vaccines -6 Study G, Investigators, Naggie S. Effect of Higher-Dose Fluvoxamine vs Placebo on Time to Sustained Recovery in Outpatients with Mild to Moderate COVID-19: A Randomized Clinical Trial. *medRxiv*. Sep 13 2023;doi:10.1101/2023.09.12.23295424

6.	Rothman RL, Stewart TG, Mourad A, et al. Time to Sustained Recovery Among Outpatients With COVID-19 Receiving Montelukast vs Placebo: The ACTIV-6 Randomized Clinical Trial. *JAMA Netw Open*. Oct 1 2024;7(10):e2439332. doi:10.1001/jamanetworkopen.2024.39332

7.	Ma KC, Castro J, Lambrou AS, et al. Genomic Surveillance for SARS-CoV-2 Variants: Circulation of Omicron XBB and JN.1 Lineages - United States, May 2023-September 2024. *MMWR Morb Mortal Wkly Rep*. Oct 24 2024;73(42):938-945. doi:10.15585/mmwr.mm7342a1

8.	Lewnard JA, Mahale P, Malden D, et al. Immune escape and attenuated severity associated with the SARS-CoV-2 BA.2.86/JN.1 lineage. *Nat Commun*. Oct 3 2024;15(1):8550. doi:10.1038/s41467-024-52668-w

9.	Levy ME, Chilunda V, Davis RE, et al. Reduced Likelihood of Hospitalization with the JN.1 or HV.1 SARS-CoV-2 Variants Compared to the EG.5 Variant. *J Infect Dis*. Jul 19 2024;doi:10.1093/infdis/jiae364

10.	Li P, Faraone JN, Hsu CC, et al. Neutralization escape, infectivity, and membrane fusion of JN.1-derived SARS-CoV-2 SLip, FLiRT, and KP.2 variants. *Cell Rep*. Aug 27 2024;43(8):114520. doi:10.1016/j.celrep.2024.114520

11.	Kumar P, Jayan J, Sharma RK, et al. The emerging challenge of FLiRT variants: KP.1.1 and KP.2 in the global pandemic landscape. *QJM*. Jul 1 2024;117(7):485-487. doi:10.1093/qjmed/hcae102

12.	Qu P, Evans JP, Faraone JN, et al. Enhanced neutralization resistance of SARS-CoV-2 Omicron subvariants BQ.1, BQ.1.1, BA.4.6, BF.7, and BA.2.75.2. *Cell Host Microbe*. Jan 11 2023;31(1):9-17 e3. doi:10.1016/j.chom.2022.11.012

13.	Wang Q, Mellis IA, Ho J, et al. Recurrent SARS-CoV-2 spike mutations confer growth advantages to select JN.1 sublineages. *Emerg Microbes Infect*. Dec 2024;13(1):2402880. doi:10.1080/22221751.2024.2402880
